# SARS-CoV-2 sculpts the immune system to induce sustained virus-specific naïve-like and memory B cell responses

**DOI:** 10.1101/2021.04.29.21256002

**Authors:** Leire de Campos-Mata, Sonia Tejedor Vaquero, Roser Tachó-Piñot, Janet Piñero, Emilie K. Grasset, Itziar Arrieta Aldea, Natalia Rodrigo Melero, Carlo Carolis, Juan P. Horcajada, Andrea Cerutti, Judit Villar-García, Giuliana Magri

**Affiliations:** Translational Clinical Research Program, Institut Hospital del Mar d’Investigacions Mèdiques (IMIM), 08003 Barcelona, Spain; Research Programme on Biomedical Informatics (GRIB), Hospital del Mar Medical Research Institute (IMIM), Department of Experimental and Health Sciences, Pompeu Fabra University (UPF), Barcelona, Spain; Department of Medicine, Immunology Institute, Icahn School of Medicine at Mount Sinai, New York, NY 10029, USA; Department of Infectious Diseases, Hospital Del Mar, Institut Hospital del Mar d’Investigacions Mèdiques (IMIM), 08003, Barcelona, Spain; Centre for Genomic Regulation (CRG), The Barcelona Institute of Science and Technology, Dr. Aiguader, 88, 08004 Barcelona; Catalan Institute for Research and Advanced Studies (ICREA), 08003 Barcelona, Spain

## Abstract

SARS-CoV-2 infection induces virus-reactive memory B cells expressing unmutated antibodies, which hints at their emergence from naïve B cells. Yet, the dynamics of virus-specific naïve B cells and their impact on immunity and immunopathology remain unclear. Here, we longitudinally studied moderate to severe COVID-19 patients to dissect SARS-CoV-2-specific B cell responses overtime. We found a broad virus-specific antibody response during acute infection, which evolved into an IgG1-dominated response during convalescence. Acute infection was associated with increased mature B cell progenitors in the circulation and the unexpected expansion of virus-targeting naïve-like B cells that further augmented during convalescence together with virus-specific memory B cells. In addition to a transitory increase in tissue-homing CXCR3^+^ plasmablasts and extrafollicular memory B cells, most COVID-19 patients showed persistent activation of CD4^+^ and CD8^+^ T cells along with transient or long-lasting changes of key innate immune cells. Remarkably, virus-specific antibodies and the frequency of naïve B cells were among the major variables defining distinct immune signatures associated with disease severity and inflammation. Aside from providing new insights into the complexity of the immune response to SARS-CoV-2, our findings indicate that the de novo recruitment of mature B cell precursors into the periphery may be central to the induction of antiviral immunity.

## Introduction

To date, the rapidly spreading severe acute respiratory syndrome coronavirus 2 (SARS-CoV-2) has infected around 142 million people, resulting in more than three million deaths worldwide^1^. Infection with SARS-CoV-2 causes the coronavirus disease 2019 (COVID-19), which is characterized by a wide variety of clinical manifestations that range from asymptomatic to acute respiratory distress syndrome (ARDS), multi organ failure and death^2^. Although such diversity in disease pathogenesis is partially explained by the patient’s comorbidities as well as genetic and socio-demographic factors, severe manifestations of the disease are strongly associated with dysregulated immune responses^3,4^. Immune dysregulation in severe COVID-19 patients is characterized by delayed and impaired type I interferon responses that associate with failure to control primary infection^5,6^. The resulting aberrant activation of innate immune cells leads to an exacerbated release of pro-inflammatory cytokines, causing systemic inflammation and tissue damage^7^. Interestingly, interferon signaling and hyper inflammation may associate with autoimmunity. Indeed, severe COVID-19 patients develop autoantibodies against immunomodulatory proteins, including antibodies against type I interferon^8–10^.

Besides playing a role in immunopathogenesis, the host immune response is a major determinant of recovery and immune protection through the development of durable SARS-CoV-2-specific T and B cell responses. Several studies have documented the early activation of CD4 and CD8 T cells following SARS-CoV-2 infection as well as the generation of durable virus-specific T cell responses required for immune protection^11–14^. In the early response to SARS-CoV-2, infected individuals generate antibodies against the viral nucleocapsid (NP) and spike (S) proteins. Around 90% of COVID-19 patients produce detectable neutralizing antibody responses against the receptor binding domain (RBD) of the viral S protein, which persist for up to 8 months^15–17^. Early humoral responses are driven by the transient expansion of antibody-secreting plasmablasts. During convalescence, humoral memory is sustained by somatically-mutated memory switched B cells and long-lived plasma cells^14,16,18,19^. Of note, recent studies identified convergent antibody responses to SARS-CoV-2 by B cells with preferential immunoglobulin heavy chain variable-joining (IGHV-J) gene usage and minimal somatic hypermutation^18,20–25^. These findings suggest that humoral protection involves SARS-CoV-2 recognition by naïve B cells with little or no antigen-driven affinity maturation required^21,26^.

In spite of our growing understanding of SARS-CoV-2 infection, both kinetics and composition of virus-specific B cell responses remain poorly understood. In particular, the dynamics of virus-reactive naïve B cells and their role in immune protection and immunopathology are unclear. In addition, the temporal trajectories of innate and adaptive immune responses to SARS-CoV-2 and their functional relationship remain elusive. A better understanding of these facets of SARS-CoV-2 infection may help in the evaluation of the protective effects afforded by individual vaccination programs.

Here, we longitudinally profiled global and virus-specific B cell responses from a cohort of moderate to severe COVID-19 patients at different stages of SARS-CoV-2 infection. We also explored the relationship of B cell responses to SARS-CoV-2 with the activation of effector and regulatory cells from the innate or adaptive immune system. We identified specific properties of immune response dynamics from COVID-19 patients as well as unique immune trajectories that associate with disease severity and inflammation. We also found some evidence of a novel mechanism adopted by the host immune system to fight SARS-CoV-2. This mechanism may involve the continuous peripheral recruitment of early mature B cell precursors to enhance viral recognition by the naïve B cell repertoire.

## Results

### SARS-CoV-2 triggers a broad antibody response in the early stage of infection and a robust and long-lasting RBD-specific IgG1 response

To dissect the dynamics of humoral immune responses to SARS-CoV-2, we collected blood and serum samples from a cohort of 25 hospitalized COVID-19 patients in the acute phase of infection (COVT1). Of these patients, 24% (*n* = 6; darker symbols in Figures) were admitted to intensive care unit (ICU). On average, COVT1 samples were collected 11 days post-symptom onset (PSO). A follow-up blood sample was drawn from 20 of 25 patients (COVT2), including 4 ICU patients, averaging 70 days PSO (**Fig. 1A**). Blood and serum samples were also collected from healthy controls (*n* = 21). Aside from being a major risk factor for fatal outcome in COVID-19 patients, age profoundly influences the immune system of healthy individuals. To minimize these age-related effects, we mostly selected patients younger than 65 (median age = 51) and included age-matched healthy controls (median age = 50). A summary of demographic and clinical data of all the individuals included in this study is provided (**Table S1)**.

**Fig. 1.**
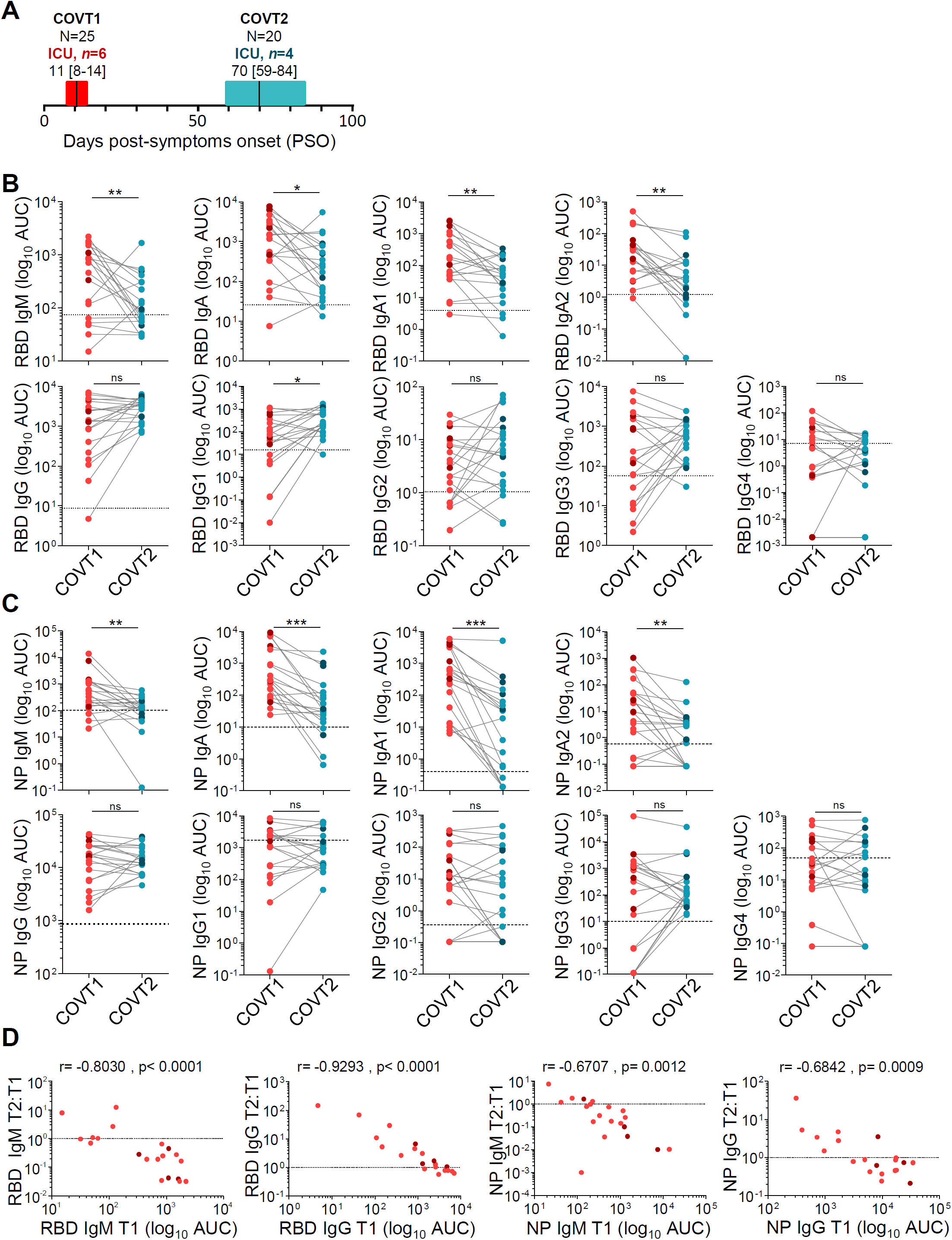
SARS-CoV-2 induces a broad antibody response in the early stage of infection and a long-lasting IgG response. (**A**) Schematic diagram of the study timeline and cohort characteristics. Range of days post-symptom onset (PSO) is indicated with a box and median is indicated with a line for COVID-19 patients in the acute (COVT1) and convalescent (COVT2) phase of infection. (**B**) Area under the curve (AUC) for each of the RBD-specific and (**C**) NP-specific antibody classes and subclasses analyzed from COVT1 and COVT2 sera samples. Sera from healthy controls (HCs) were analyzed in parallel to establish negative threshold values defined as the HC AUC mean plus 2 times the standard deviation of the mean. Data are presented as paired individual dots. Dashed line indicates negative threshold. Wilcoxon matched pairs test (*P < 0.05, **P < 0.01, and ***P < 0.001). (**D**) Relative change in antibody levels between COVT1 and COVT2 plotted against the corresponding antibody levels at COVT1. r stands for Spearman’s rank-order correlation. (**A-D**) Dark-colored dots show ICU patients. COVT1, *n*=20; COVT2, *n*=20.

To elucidate whether COVID-19 patients can mount a broad and long-lasting humoral response to SARS-CoV-2, we performed isotype-specific (IgM, IgA and IgG) and subtype-specific (IgA1, IgA2, IgG1, IgG2, IgG3 and IgG4) enzyme-linked immunosorbent assays (ELISAs) that quantified antibody responses to the receptor-binding domain (RBD) of the spike protein or nucleocapsid protein (NP) from SARS-CoV-2. Compared to healthy controls, sera from COVID-19 patients in acute infection (COVT1) and convalescence (COVT2) showed significantly higher titers of all RBD- and NP-specific antibody classes and subclasses analyzed (**Fig. S1**). According to our longitudinal study and as previously reported^18,27^, RBD- and NP-specific IgM and IgA titers significantly decreased two months PSO (**Fig. 1B, C**), presumably because they mainly originated from short-lived plasmablasts (PBs). Interestingly, both IgA1 and IgA2 subclasses contributed to the decline of virus-specific IgA, suggesting a limited involvement of mucosal immunity during convalescence. Unlike RBD- and NP-specific IgM and IgA, virus-specific IgG titers were maintained over the first two months PSO (**Fig. 1B, C**). Remarkably, RBD-specific IgG1 was the only IgG subclass that significantly increased overtime (**Fig. 1B**).

We further studied the dynamics of the humoral response in COVID-19 throughout the study’s time course and found that the magnitude of relative changes in virus-specific IgM or IgG titers directly correlated with their initial levels at COVT1. Indeed, patients with initial higher IgM or lower IgG titers showed greater relative decrease or increase, respectively (**Fig. 1D**), similarly to what has been previously reported for convalescent individuals at later time points^18^. Thus, SARS-CoV-2 infection triggers a broad antibody response in terms of antigen-specificity and antibody isotypes in early stages and an expansion of an RBD-specific and IgG1-dominated response during convalescence.

### COVID-19 is associated with transitory expansion of circulating plasmablasts and naïve B cells during the acute phase of infection

To further explore the dynamics of the B cell response to SARS-CoV-2, we analyzed circulating B cells from healthy controls and COVID-19 patients in the acute and convalescent phase of infection by high-dimensional spectral flow cytometry (**Table S2**). As reported by others^28^, the frequency of CD19^+^ B cells within live peripheral blood mononuclear cells (PBMCs) was significantly higher in COVT1 compared to healthy controls or COVT2 (**Fig. 2A**). T-distributed stochastic neighbour embedding (tSNE) defined the major B cell populations (**Fig. 2B**), the contribution of each group of samples to these populations (**Fig. 2C**), and the differential expression of multiple surface B cell proteins within these populations (**Fig. 2D**). In parallel, we queried the data by traditional gating (**Fig. S2A, C**). This analysis revealed that, for most COVID-19 patients, the acute response to SARS-CoV-2 was dominated by a transitory expansion of CD38^++^CD10−CD27^high^ plasma cells (PCs; **Fig. 2B-F**), the majority of which expressed HLA-DR^+^ (**Fig. 2G**), an antigen-presenting protein typically expressed by newly generated short-lived PBs^29^. Next, the analysis of antibody classes and subclasses confirmed the induction of unswitched as well as IgA1 or IgG class-switched PCs soon after infection (**Fig. 2H** and **Fig. S2B**). We then assessed the homing potential of these PCs through the analysis of CXCR3 and CXCR4 chemokine receptors, which guide PCs to inflamed tissues or bone marrow, respectively^30^. We found that the majority of PCs from COVT1 had a CXCR3^+^CXCR4^-^ phenotype, suggesting their targeted migration into inflamed tissues (**Fig. 2I, J**). Compared to healthy controls and COVT2, COVT1 samples also showed a significantly increased proportion of CXCR3^+^CXCR4^+^ PCs (**Fig. 2I, K**).

**Fig. 2.**
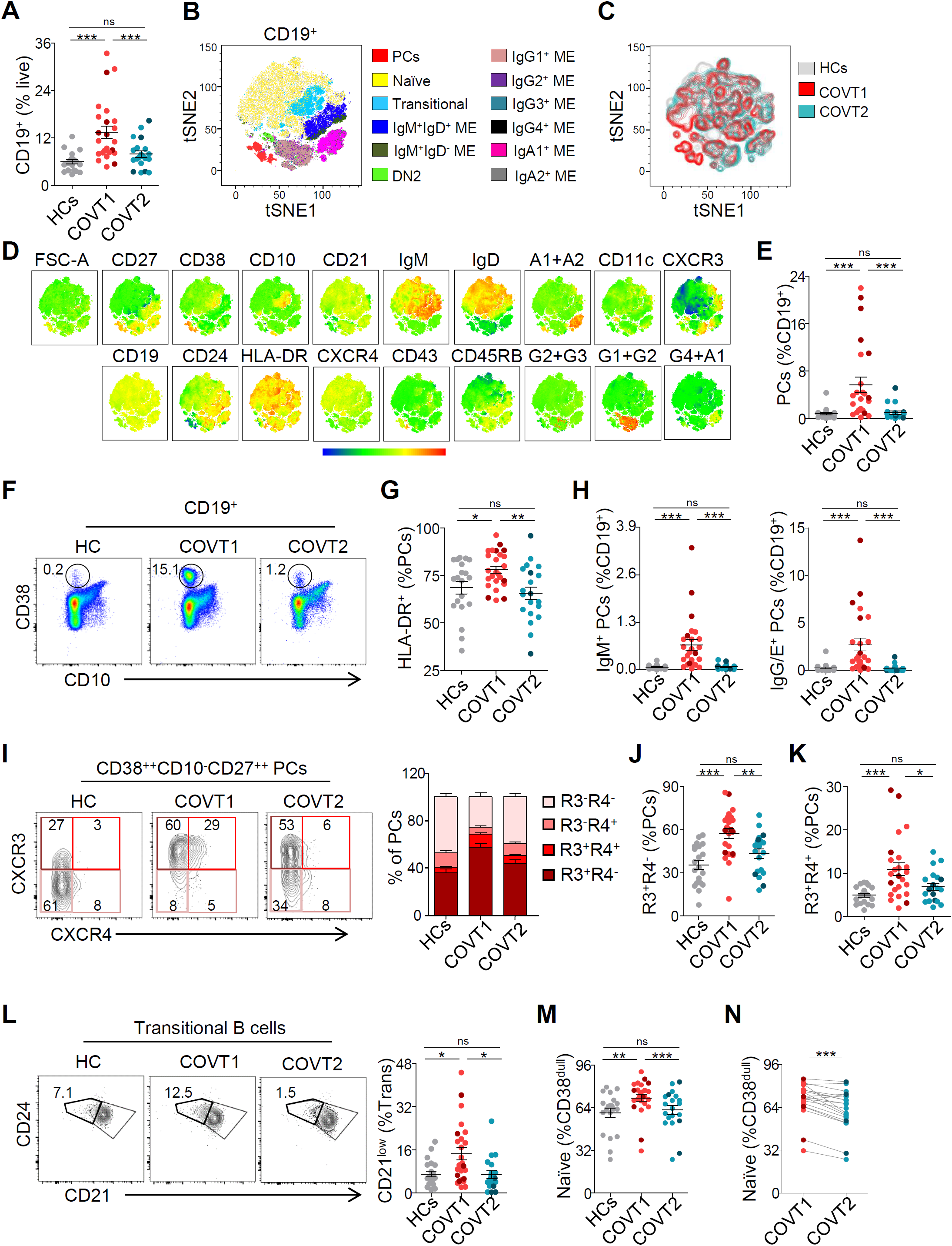
Deep profiling of B cell subsets in COVID-19 patients reveals transitory expansion of circulating plasmablasts, immature transitional and naïve B cells during the acute phase of infection. (**A**) Frequency of CD19^+^ B cells from live peripheral blood mononuclear cells (PBMCs) in healthy controls (HCs) and COVID-19 patients at T1 (COVT1) and at T2 (COVT2). (**B**) Merged tSNE projection of CD19^+^ B cells for HCs (*n*=11), COVT1 (*n*=16) and COVT2 (*n*=16) samples concatenated and overlaid, with main B cell populations indicated by color. PCs, plasma cells. ME, memory. DN2, CD38^dull^CD10^−^IgD^-^CD27^-^CD21^-^CD11c^+^ double negative type 2. (**C**) Merged tSNE projection of CD19^+^ B cells with each sample group indicated by color. (**D**) tSNE projections of expression of the indicated cell surface markers. (**E**) Frequency of CD38^++^CD10^−^CD27^+^ PCs from total CD19^+^ B cells and (**F**) representative dot plots in HCs, COVT1 and COVT2. Numbers indicate percentages in the drawn gates. (**G**) Frequency of HLA-DR^+^ plasmablasts (PBs) from total circulating CD38^++^CD10^−^CD27^+^ cells in HCs, COVT1 and COVT2. (**H**) Frequencies of IgM^+^ PCs (left) and IgG/E^+^ PCs (right) from total CD19^+^ B cells. (**I**) Representative flow cytometry plots (left) and relative percentage (right) of CXCR3^-^CXCR4^-^ (R3^-^R4^-^), CXCR3^-^CXCR4^+^ (R3^-^R4^+^), CXCR3^+^CXCR4^+^ (R3^+^R4^+^), CXCR3^+^CXCR4^-^ (R3^+^R4^-^) PCs measured in each group of samples. Numbers indicate percentages in the drawn gates. (**J**) Frequency of CXCR3^+^CXCR4^-^ cells within total circulating PCs. (**K**) Frequency of CXCR3^+^CXCR4^+^ cells within total circulating PCs. (**L**) Representative flow cytometry plots (left) and frequency (right) of immature cells (CD38^int^CD10^+^IgD^+^CD27^-^CD21^low^) from total transitional B cells, gated with thick black line. Numbers indicate percentages in the drawn gates. (**M**) Frequency of naïve B cells (non-PC CD19^+^CD27^-^IgD^+^) from CD19^+^CD38^dull^ B cells in each group of samples. (**N**) Paired analysis of naïve B cell frequency in COVT1 (*n*=20) and COVT2 (*n*=20). Data are presented as individual dots. Bars represent mean ± SEM. Dark-colored dots show ICU patients. Two-tailed Mann-Whitney U test was performed to compare HCs with COVT1 and HCs with COVT2. Wilcoxon matched pairs test was performed to compare COVT1 with COVT2 (*P < 0.05, **P < 0.01, and ***P < 0.001). Unless mentioned otherwise, HCs, *n*=19; COVT1, *n*=25; COVT2, *n*=20.

The tSNE projections also highlighted prominent differences in the transitional and naïve B cell compartments among healthy controls, COVT1 and COVT2 (**Fig. 2B-D**). Using conventional gating strategies (**Fig. S2C**) and pairwise longitudinal comparisons, we identified an increased proportion of immature CD21^low^ transitional B cells (**Fig. 2L**) as well as a highly significant transient expansion of naïve B cells during the acute phase of infection (**Fig. 2M, N**). Furthermore, our data revealed a positive correlation between the frequency of naïve B cells and the frequency of CD19^+^ B cells (**Fig. S2D**), which implies that naïve B cell expansion is responsible for the increased frequency of CD19^+^ B cells. These changes likely reflect an increased inflammation-dependent mobilization of developing B cells from the bone marrow into the periphery, a phenomenon also observed in other infections^31^. Compared to similar cells from healthy controls, naïve B cells from COVT1 exhibited significantly lower expression of CD21, HLA-DR, IgD and the chemokine receptors CCR6, CXCR5 and CXCR3 but not CCR7, presumably due to their activation by signals from the inflammatory environment (**Fig. S2E-K**). Thus, aside from promoting a transient induction of CXCR3^+^ unswitched and class-switched PBs, SARS-CoV-2 may induce the mobilization of precursors of mature B cells together with the transient expansion and activation of naïve B cells.

### SARS-CoV-2 infection drives transient increases of extrafollicular memory switched B cells and long-lasting expansion of IgG1 memory B cells

To gain insight into the dynamics of B cell memory (ME) responses to SARS-CoV-2, we analyzed the frequency of different antigen-experienced B cell populations in healthy controls and COVID-19 patients during acute infection and convalescence. First, we found that the frequency of ME B cells within total CD19^+^ B cells was significantly lower in acute COVT1 compared to COVT2 and healthy controls (**Fig. S3A** and **Fig. 3A**), presumably due to the transient expansion of newly formed PBs and naïve B cells. Unlike PCs but similar to naïve B cells, the frequency of CXCR3^+^ ME B cells diminished during the acute phase of infection (**Fig. 3B**), suggesting a possible impairment in their recruitment into inflamed tissues and lymphoid follicles^32^. Within the ME B cell compartment, we observed a significant and persistent reduction in the proportion of IgM^+^IgD^+^CD27^+^ ME unswitched B cells in COVID-19 patients (**Fig. S3A** and **Fig. 3C**) and a progressive expansion of IgG1-expressing ME B cells (**Fig. 3D**) within the memory compartment. Interestingly, the frequency of unswitched ME B cells positively correlated with the serum concentration of inflammatory marker ferritin (**Fig. 3E**) as well as with NP-specific IgM titers (**Fig. 3F**). Moreover, COVID-19 patients with higher plasma concentration of ferritin and other markers of inflammation (data not shown) also showed a lower frequency of IgG1 ME B cells during acute infection (**Fig. 3G**). Remarkably, ME B cells expressing other IgG subclasses did not increase after infection (**Fig. S3B**). These results may reflect the consolidation of efficient anti-viral IgG1-mediated immunity in less severe patients.

**Fig. 3.**
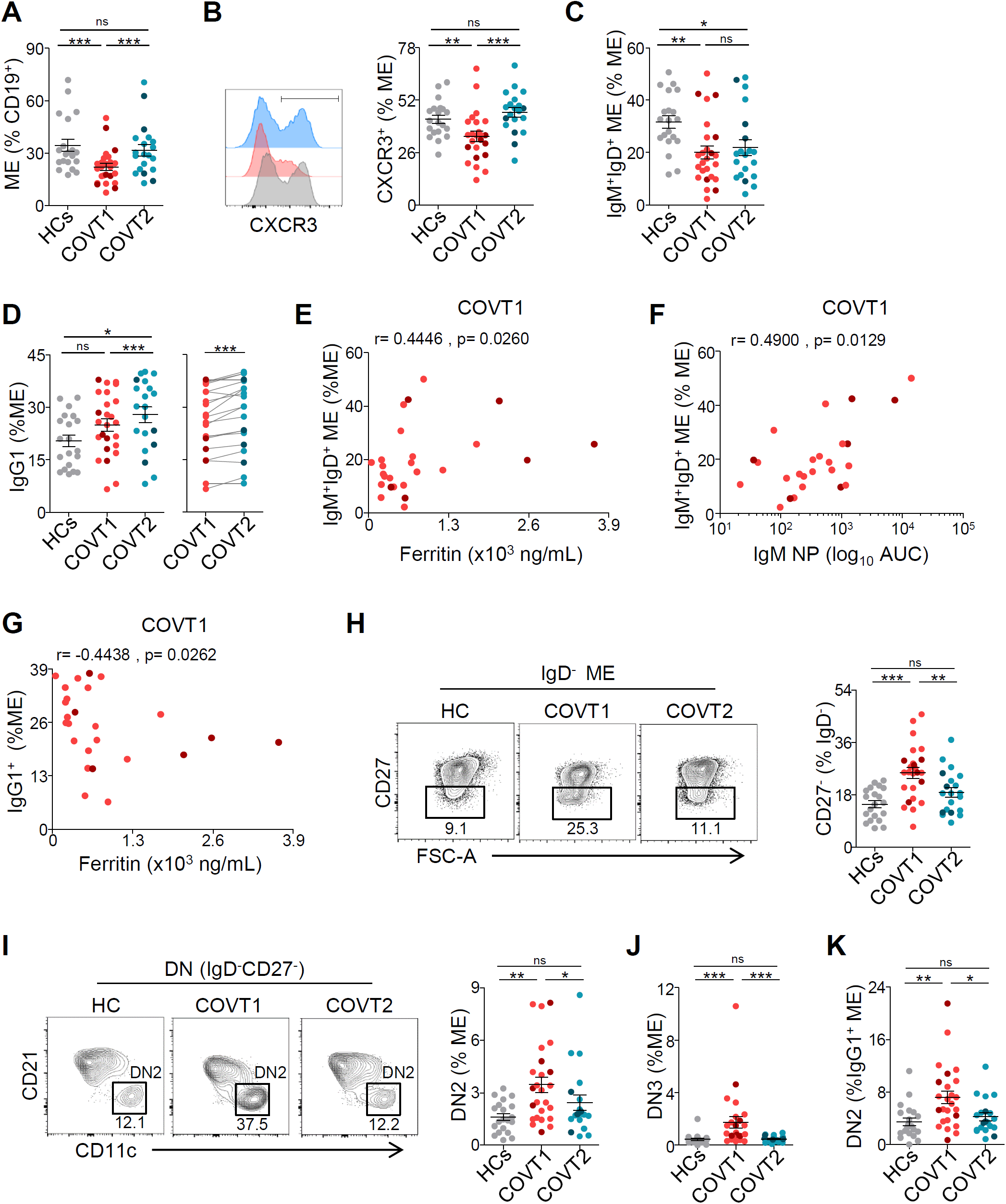
COVID-19 is associated with temporary expansion of extrafollicular switched memory B cells, long-lasting contraction of IgM^+^IgD^+^CD27^+^ memory B cells and late increase in IgG1^+^ memory B cells. (**A**) Frequency of memory (ME) B cells defined as depicted in **Fig. S3** from total CD19^+^ B cells in healthy controls (HCs), COVT1 and COVT2. (**B**) Representative flow cytometry histogram and frequency of CXCR3^+^ cells from total ME B cells in HCs, COVT1 and COVT2. (**C**) Frequency of CD38^dull^CD10^−^IgM^+^IgD^+^CD27^+^ ME B cells within total ME B cells in HCs, COVT1 and COVT2. (**D**) Frequency of IgG1^+^ ME B cells within total ME B cells in HCs, COVT1 and COVT2 (left) and paired analysis in COVT1 (*n*=20) and COVT2 (*n*=20, right). (**E**) Spearman correlation analysis of ferritin levels in COVT1 plotted against CD38^dull^CD10^−^ IgM^+^IgD^+^CD27^+^ ME B cells within total ME B cells in COVT1. (**F**) Spearman correlation analysis of NP-specific IgM antibody titers in COVT1 plotted against CD38^dull^CD10^−^ IgM^+^IgD^+^CD27^+^ ME B cells within total ME B cells in COVT1. (**G**) Spearman correlation analysis of IgG1 ME B cells within total ME B cells in COVT1 plotted against ferritin levels in COVT1. (**E-G**) r stands for Spearman’s rank-order correlation. (**H**) Representative flow cytometry plots and frequency of DN memory B cells defined as CD38^dull^CD10^−^IgD^-^CD27^-^ out of total IgD^-^ ME B cells. Numbers indicate percentages in the drawn gates. (**I**) Representative flow cytometry plots and frequency of extrafollicular DN2 ME B cells defined as CD38^dull^CD10^−^IgD^-^CD27^-^CD21^-^CD11c^+^ out of total ME B cells. Numbers indicate percentages in the drawn gates. (**J**) Frequency of DN3 ME B cells defined as CD38^dull^CD10^−^IgD^-^CD27^-^CD21^-^CD11c^-^ out of total ME B cells. (**K**) Frequency of DN2 ME B cells from total IgG1^+^ ME B cells. Data are presented as individual dots. Dark-colored dots show ICU patients. Error bars represent mean ± SEM. Two-tailed Mann-Whitney U test was performed to compare HCs with COVT1 and HCs with COVT2. Wilcoxon matched pairs test was performed to compare COVT1 with COVT2 (*P < 0.05, **P < 0.01, and ***P < 0.001). Unless mentioned otherwise, HCs, *n*=19; COVT1, *n*=25; COVT2, *n*=20.

In agreement with findings published previously^29^, we categorized IgD^-^CD27^-^ double negative (DN) ME B cells as early activated CD27^-^CD21^+^CD11c^-^ ME or DN1, extrafollicular CD27^-^CD21^-^ CD11c^+^ PB precursors or DN2, and DN3 CD27^-^CD21^-^CD11c^-^ B cells (**Fig. S3C**). The analysis of these ME B cell subsets revealed a significant and transient increase in the frequency of CD27^-^IgD^-^ DN cells in COVT1 compared to COVT2 and healthy controls (**Fig. 3H**), as reported by others^28^. Among DN B cells, DN2 cells are recognized as primed precursors of antibody-secreting cells that differentiate from newly activated naïve B cells through an extra-follicular pathway. In contrast, DN3 has unknown origin and function^28^. First, we quantified the frequency of DN2 cells within different ME B cells subsets distinguished on the basis of their expressed isotype or subclass. We found a significantly increased proportion of DN2 in ME B cells expressing IgG3, IgG1 and, to lesser extent, IgA1 (**Fig. S3D**). The frequency of extrafollicular ME DN2 B cells and DN3 cells were higher in COVT1 compared to COVT2 and healthy controls (**Fig. 3I, J**). Interestingly, the most significant increase in DN2 was observed within the IgG1^+^ ME B cell subset (**Fig. 3K**). Thus, SARS-CoV-2 infection induces the transient expansion of extrafollicular IgG1^+^ ME B cells, which is followed by a long-lasting IgG1^+^ ME B cell response.

### SARS-CoV-2 induces RBD-specific PBs during acute infection followed by sustained expansion of circulating RBD-specific naïve and switched memory B cells in convalescence

Next, a fluorescently labeled recombinant RBD probe was used to identify SARS-CoV-2-specific B cells capable of producing potentially neutralizing antibodies (**Fig. S4A**). We confirmed the specificity of our assay by comparing our results to those obtained using a double discrimination strategy via the inclusion of two fluorescently labeled RBD probes (**Fig. S4B**). Antigen-binding CD19^+^ B cells were further characterized according to the expression of CD27, CD21, CD11c, HLADR, IgM, IgD, IgA and the Ig light chain λ (**Fig. S4C** and **Table S3**). In agreement with published studies^14,17,33^, SARS-CoV-2 infection induced a rapid increase in the frequency of RBD-specific CD19^+^ B cells at T1 that persisted 2 months after viral exposure (**Fig. 4A**). Interestingly, the phenotypic characterization of these antigen-binding B cells revealed significant changes in virus-specific B cell populations between the acute and convalescent phase of infection (**Fig. 4B** and **Fig. S4D**). Consistent with the observed expansion of PCs, COVT1 showed an increased frequency of RBD-specific CD27^high^CD21^-^ PCs compared to healthy controls and COVT2 (**Fig. 4C** and data not shown). Remarkably, virtually 100% of these RBD-specific cells were newly generated HLADR^+^ PBs (**Fig. 4D**).

**Fig. 4.**
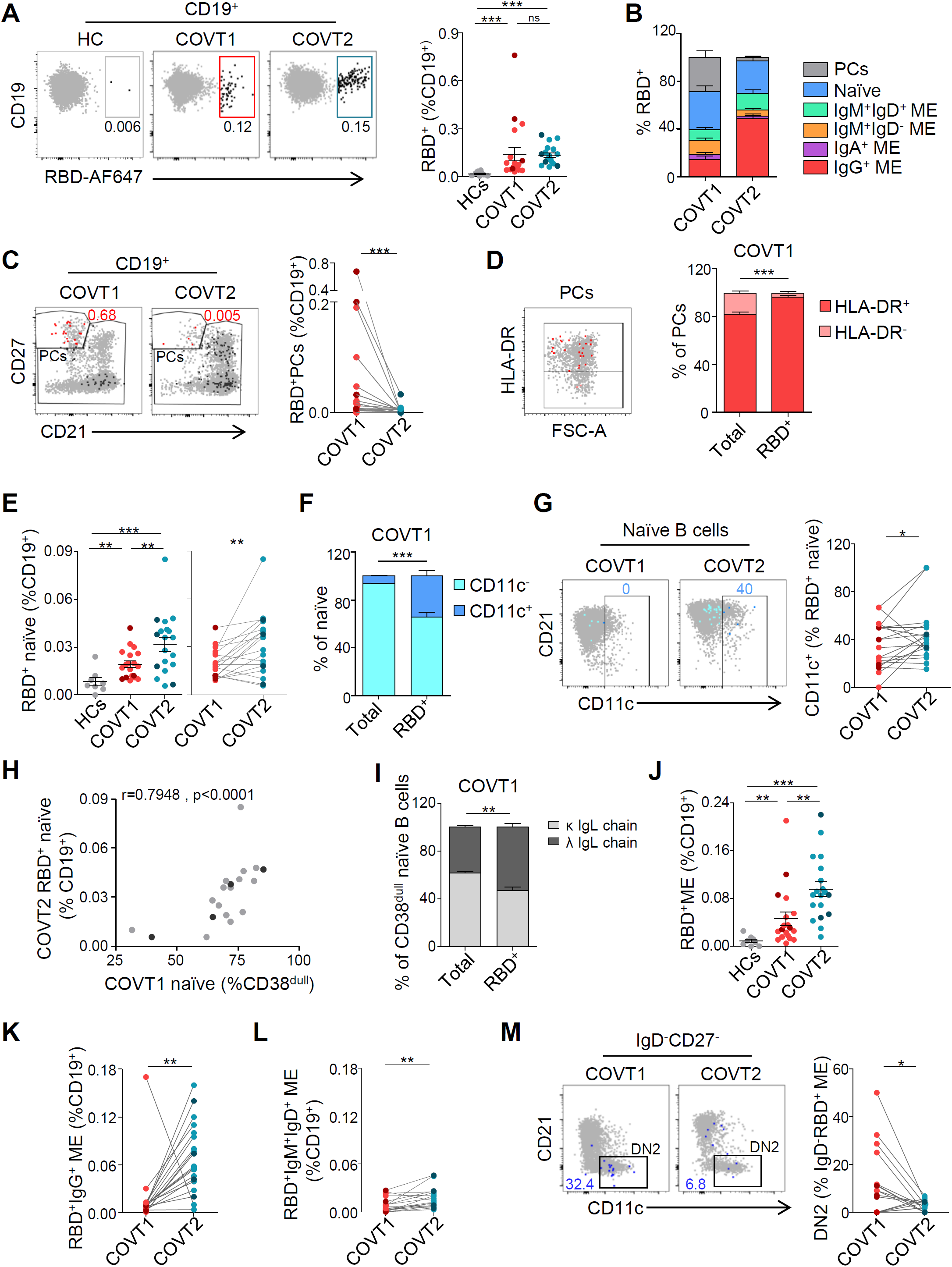
SARS-CoV-2 induces a temporal enrichment of RBD-specific plasmablasts in the early stage of infection and a sustained expansion of RBD-specific naïve and memory B cell subsets. (**A**) Representative flow cytometry staining of CD19^+^ RBD-specific B cells (left) and frequency of RBD^+^ cells within total CD19^+^ B cells (right) from each group of samples using a fluorescently labeled RBD probe. Numbers indicate the percentage of RBD-specific cells within total CD19^+^ B cells. Black large dots represent RBD-specific B cells. (**B**) Relative percentage of B cell subsets within total RBD^+^ CD19^+^ B cells in healthy controls (HCs), COVT1 and COVT2. (**C**) Representative flow cytometry plots and paired analysis of the frequency of RBD-specific PCs (CD19^+^RBD^+^CD27^++^CD21^-^) within total CD19^+^ B cells. In the flow cytometry plot, red large dots represent RBD-specific PCs and numbers indicate percentage of RBD-specific PCs for the drawn sample. (**D**) Representative flow cytometry plot and relative percentage of total and RBD-specific PCs expressing HLA-DR in COVT1. In the flow cytometry plot, red large dots represent RBD-specific PCs expressing HLA-DR. (**E**) Frequency and paired analysis of RBD-specific naïve B cells (non-PC CD19^+^RBD^+^CD27^-^ IgD^+^) within total CD19^+^ in HCs, COVT1 and COVT2. (**F**) Relative percentage of total or RBD-specific naïve B cells expressing CD11c in COVT1 samples. (**G**) Representative flow cytometry plots and paired analysis of the frequency of CD11c^+^ cells from RBD-specific naïve B cells. Numbers indicate percentage of RBD-specific CD11c^+^ naïve cells in the drawn gates. (**H**) Spearman correlation analysis of CD38^dull^ naïve B cells in COVT1 and RBD^+^ naïve B cells within CD19^+^ cells in COVT2. r stands for Spearman’s rank-order correlation. (**I**) Relative percentage of total and RBD^+^ naïve B cells expressing lambda or kappa light chains in COVT1. (**J**) Frequency of RBD-specific ME B cells defined as non-PC CD19^+^RBD^+^IgD^-^ from total CD19^+^ B cells in HCs, COVT1 and COVT2. (**K**) Paired analysis of the frequency of RBD-specific IgG^+^ ME B cells defined as IgA^-^IgM^-^IgD^-^ within total CD19^+^ in COVT1 and COVT2. (**L**) Paired analysis of the frequency of RBD-specific IgM^+^IgD^+^ ME B cells defined as non-PC CD19^+^CD27^+^IgM^+^IgD^+^ within total CD19^+^ in COVT1 and COVT2. (**M**) Representative flow cytometry plots and paired analysis of the frequency of DN2 ME B cells (non-PC CD19^+^RBD^+^CD27^-^IgD^-^CD11c^+^) from total IgD^-^ RBD^+^ ME B cells. Numbers indicate percentages of RBD-specific DN2 ME in the drawn gates. Dark-colored dots show ICU patients. Bars represent mean ± SEM. Two-tailed Mann-Whitney U test was performed to compare HCs with COVT1 and HCs with COVT2. Wilcoxon matched pairs test was performed to compare COVT1 with COVT2 (*P < 0.05, **P < 0.01, and ***P < 0.001). HCs, *n*=11; COVT1, *n*=19; COVT2, *n*=19. (**B, D, F, I)** Data are presented as mean ± SEM.

In addition, RBD-targeting IgM^+^IgD^+^CD27^-^ naïve B cells increased progressively in COVID-19 patients (**Fig. 4E**). These cells were enriched in CD11c^+^ early activated B cells in COVT1 and even more in COVT2 (**Fig. 4F-G**). Of note, the frequency of RBD-specific naïve B cells during convalescence strongly correlated with the frequency of total naïve B cells in the acute phase of infection (**Fig. 4H**), suggesting that the early mobilization of developing B cells to the periphery and the expansion of naïve B cells could contribute to humoral defense by increasing germline RBD-specific B cells. To elucidate the mechanisms leading to the sustained enrichment of RBD-targeting naïve B cells, we measured the plasma concentration of IL-7, a cytokine involved in B and T lymphopoiesis^34^. Compared to healthy controls, COVID-19 patients showed more IL-7 during acute infection (**Fig. S4E**) and its concentration correlated with the frequency of RBD-specific naïve B cells in COVT2 (**Fig. S4F**), supporting a direct role of this cytokine in enhancing the expansion and recruitment of relatively immature transitional and naïve B cells with useful specificities. Additionally, RBD-specific naïve B cells expressed more Igλ compared to total naïve B cells (**Fig. 4I**). Given that Igλ light chain has a more varied structural repertoire than Igκ^35^, Igλ enrichment may provide the host with the advantage of developing a broader humoral response to SARS-CoV-2.

We then analyzed whether RBD-specific ME B cells were induced and maintained throughout the study’s time course. The number of RBD^+^ ME B cells in COVT1 was significantly greater than in healthy controls and increased even further in COVT2 (**Fig. 4J**). Similarly, there was a sustained increase in the number of RBD-specific IgG (IgM^-^ IgD^-^IgA^-^) class-switched ME and, to a lesser extent, in RBD-specific IgM^+^IgD^+^CD27^+^ unswitched ME B cells (**Fig. 4K-L**). The RBD-specific ME compartment included only a few IgA^+^ and unswitched IgM^+^IgD^-^ ME cells and these subsets did not increase overtime (**Fig. S4G**). Consistent with our characterization of B cell subsets, RBD-specific extrafollicular ME B cells, or DN2, expanded during the acute phase of infection but their frequency dramatically decreased in convalescence (**Fig. 4M**).

To assess the persistency of virus-specific ME B cell responses, RBD-targeting B cell subsets were analyzed by flow cytometry in a small cohort of non-hospitalized convalescent individuals three months (COVT3) and six months (COVT6) PSO (**Fig. S5**). In agreement with previous reports^14,17,18^, RBD-specific B cells persisted up to 6 months after infection (**Fig. S5A**) and mainly consisted of IgG^+^ ME B cells (**Fig. S5B**). Accordingly, RBD-specific IgG and, to a lesser extent, IgA were significantly higher in sera from COVT3 and COVT6 compared to healthy controls (**Fig. S5C**). Thus, SARS-CoV-2 infection promotes a rapid and transient induction of RBD-specific PBs and extrafollicular ME B cells as well as a long-lasting expansion of RBD-targeting activated naïve B cells and IgG class-switched ME B cells.

### COVID-19 is associated with a sustained activation of CD4^+^ and CD8^+^ T cells

To further characterize the dynamics of immune responses over time in COVID-19 patients, we simultaneously analyzed by multicolor spectral flow cytometry the phenotypic landscape of other circulating lymphoid and myeloid populations using an in-house developed 23-color antibody panel (**Table S4**). The projection of the data for CD19^-^CD3^+^ cells in tSNE space allowed the definition of the main T cell subsets, the contribution of each group of samples to T cell populations, and the expression patterns of relevant T cell surface proteins (**Fig. 5A-C**). In parallel, we queried the data by traditional gating (**Fig. S6**). As reported previously^12,36^, the acute phase of infection was characterized by a striking reduction in the frequency of CD3^+^ T cells (**Fig. 5D**), which was driven by a decrease in the frequency of CD4^+^ and, to an even larger extent, CD8^+^ T cells (**Fig. S7A, B**). Compared to healthy controls, COVT2 convalescent individuals still had a significantly decreased frequency of CD3^+^ cells (**Fig. 5D)** and in particular of CD4^+^ T helper cells (**Fig. S7A**), which suggested their persistent loss from the periphery. We then evaluated how SARS-CoV-2 infection associated with temporary or long-lasting perturbations in both CD4 and CD8 T cell compartments. Consistent with findings published earlier^12^, the frequency of naïve T cells within total CD4^+^ T cells in acutely infected patients was significantly lower compared to age-matched healthy controls and further declined during convalescence (**Fig. 5E**). This reduction was associated with a prominent expansion of activated CD38^+^HLA-DR^+^ CD4^+^ T cells during the acute and, to a lesser extent, convalescent phase of infection (**Fig. 5F**), as observed in other viral infections^37,38^.

**Fig. 5.**
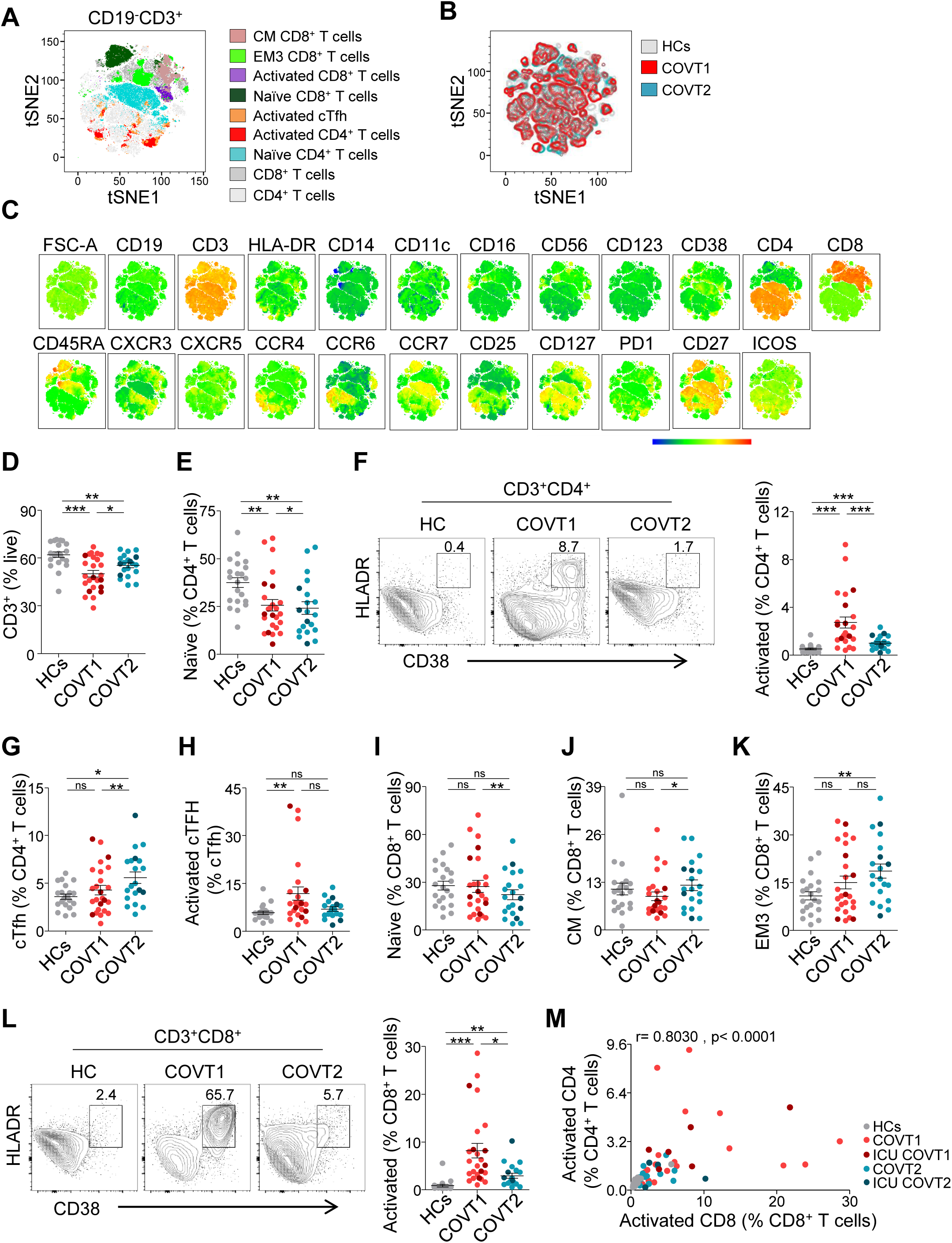
COVID-19 is associated with long-lasting contraction of naïve T cell compartment, CD4^+^ and CD8^+^ T cell activation, and expansion of circulating T follicular helper cells. (**A**) Merged tSNE projection of CD3^+^ cells for healthy controls (HCs; *n*=21), COVT1 (*n*=25) and COVT2 (*n*=20) donors concatenated and overlaid, with main T cell populations indicated by color. CM, central memory. EM, effector memory. cTfh, circulating T follicular helper T cells. (**B**) Merged tSNE projection of CD3^+^ T cells with each sample group indicated by color. (**C**) tSNE projections of expression of the indicated cell surface markers. (**D**) Frequency of CD3^+^ T cells within live lymphocytes. (**E**) Frequency of naïve CD4^+^ T cells (CD27^+^CD45RA^+^CCR7^+^) from total CD4^+^ T cells. (**F**) Representative flow cytometry plots (left) and frequency of CD38^+^HLA-DR^+^ activated CD4^+^ T cells within total CD4^+^ T cells. Numbers indicate percentages in the drawn gates. (**G**) Frequency of circulating cTfh cells (non-naïve CXCR5^+^PD-1^+^) within total CD4^+^ T cells. (**H**) Frequency of activated cTfh cells (CD38^+^ICOS^+^) within total cTfh cells. (**I**) Frequency of naïve CD8^+^ T cells (CD27^+^CD45RA^+^CCR7^+^) within total CD8^+^ T cells. (**J**) Frequency of CM CD8^+^ (CD27^+^CD45RA^-^CCR7^+^) and (**K**) EM3 (CD27^-^CD45RA^-^CCR7^-^) T cell subsets within total CD8^+^ T cells. (**L**) Representative flow cytometry plots and frequency of CD38^+^HLA-DR^+^ activated CD8^+^ T cells within total CD8^+^ T cells. Numbers indicate percentages in the drawn gates. (**M**) Spearman correlation analysis of activated CD4^+^ T and CD8^+^ T cells in all samples analyzed. r stands for Spearman’s rank-order correlation. (**A-M**) Data are presented as individual dots. Dark-colored dots show ICU patients. Bars represent mean ± SEM. Two-tailed Mann-Whitney U test was performed to compare HCs with COVT1 and HCs with COVT2. Wilcoxon matched pairs test was performed to compare COVT1 with COVT2 (*P < 0.05, **P < 0.01, and ***P < 0.001). HCs, *n*=21; COVT1, *n*=25; COVT2, *n*=20.

We then analyzed the frequency of circulating T follicular helper (cTFH) cells. Tfh cells are a specialized subset of CD4^+^ T cells that provide cognate help to antigen-specific B cells in the germinal center (GC) to initiate and maintain humoral immune responses^39^. These cells include a circulating counterpart, cTfh cells, which co-express PD-1 and CXCR5 as their GC-based equivalents do (**Fig. S6**). In general, cTfh cells can be used as a surrogate to evaluate the Tfh cell activity in lymphoid tissues^40^. Of note, activated CD38^+^ICOS^+^ cTfh cells likely reflect a recent exit from the GC immediately after antigen encounter^41^. The frequency of cTfh cells was significantly higher in COVT2 patients compared to healthy controls and COVT1 patients, which is consistent with a model of antigen persistency and long-lasting GC reaction previously suggested by others^22^ (**Fig. 5G**). Interestingly, recently activated CD38^+^ICOS^+^ cTfh cells were more profoundly increased during the acute phase, which probably reflects the peak of GC responses in the early phase of the infection (**Fig. 5H**).

We further explored whether SARS-CoV-2 infection promoted perturbations of other CD4^+^ helper T cells subsets, including central memory (CM) T cells, effector memory (EM) 1, 2 and 3 T cells, terminally differentiated effector memory (EMRA) T cells, T regulatory (Treg) cells as well as T helper 1 (Th1), Th1/17, Th2 and Th17 cells (**Fig. S7C**). We detected an increase in the frequency of Treg cells during the acute phase of infection and a small but significant increase in the frequencies of Th1 and Th1/17 cells in COVT2 compared to COVT1 upon pairwise comparison, whereas Th2 cells showed a decreased frequency over time (**Fig. S7C**).

Given the role of CD8^+^ T cells in viral infection and consistent with previous reports^12,36^, COVID-19 was associated with several changes in the CD8^+^ T cell compartment. We observed a significantly reduced frequency of naïve CD8^+^ T cells during convalescence as well as an expansion of CM and EM3 CD8^+^ T cells (**Fig. 5I-K**). On the other hand, our analysis showed no differences in the frequency of EM1, EM2 and EMRA CD8^+^ T cells between groups (**Fig. S7D**). We also found that activated CD38^+^HLA-DR^+^CD8^+^ T cells strongly expanded during the acute phase of infection and still persisted at a higher frequency compared to healthy controls during convalescence (**Fig. 5L**). As expected, patients with a higher increase in activated CD4^+^ T cells showed a higher frequency of activated CD8^+^ T cells as well (**Fig. 5M**). Thus, SARS-CoV-2 causes a marked T cell loss during the acute phase of infection, which is associated with long-lasting and coordinated activation of CD4^+^ and CD8^+^ T cells as well as expansion and activation of professional B cell-helping T cells.

### SARS-CoV-2 infection drives profound changes in the innate immune compartment

Acute SARS-CoV-2 infection triggers alterations in circulating innate immune cell subsets, some of which have been associated with COVID-19 severity^42^. Nevertheless, how these perturbations persist during convalescence after viral clearance is unclear. We combined global high-dimensional mapping via tSNE of CD19^-^CD3^-^ cells with traditional gating analysis to study significant alterations in the phenotype and frequency of peripheral blood innate immune cell subsets in COVT1 and COVT2 and healthy controls (**Fig. 6A-C** and **Fig. S8A**). Although the frequency of total circulating CD11c^+^HLA-DR^+^ myeloid cells among live PBMCs in COVT1 and COVT2 was comparable to that of healthy controls (**Fig. S8B**), we observed significant differences in the phenotype as well as subset composition of these myeloid cells. We initially explored the expression of the activation-induced molecule CD38 and HLA-DR on circulating myeloid cells. We found a striking induction of CD38 and a reduction in the expression of HLA-DR during acute infection (**Fig. 6D-E**). Of note, the decrease in HLA-DR may interfere with proper antigen presentation and has been directly linked to an immunosuppressive phenotype of monocytes during sepsis^5^. Indeed, HLA-DR levels strongly correlated with disease severity defined according to the patient’s oxygen requirement (**Fig. S8C**). However, these changes were only transient, as CD38 and HLA-DR returned to their homeostatic expression levels in the convalescent phase (**Fig. 6D-E**).

**Fig. 6.**
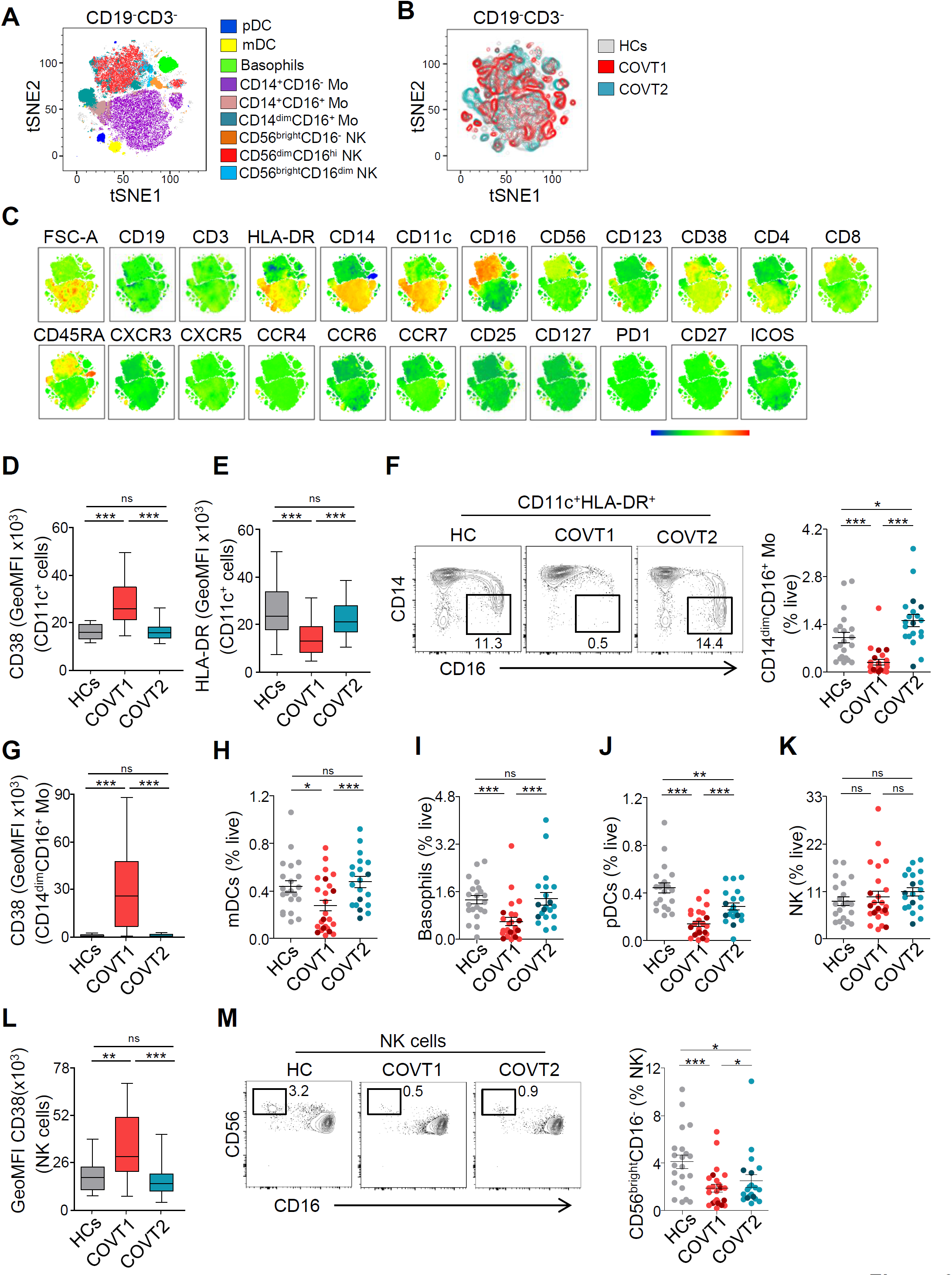
SARS-CoV-2 infection associates with transitory and long-lasting changes in the innate immune compartment. (**A**) Merged tSNE projection of CD19^-^CD3^-^ cells for healthy controls (HCs; *n*=21), COVT1 (*n*=25) and COVT2 (*n*=20) samples concatenated and overlaid, with main innate immune cell populations indicated by color. pDCs, plasmacytoid dendritic cells. mDCs, myeloid dendritic cells. Mo, monocytes. NK, natural killer cells. (**B**) Merged tSNE projection of CD19^-^CD3^-^ innate immune cells with each sample group indicated by color. (**C**) tSNE projections of indicated cell surface markers. (**D**) Geometric mean fluorescence intensity (GeoMFI) of CD38 and (**E**) HLA-DR expression in CD19^-^CD3^-^CD11c^+^ myeloid cells from HCs, COVT1 and COVT2. (**F**) Representative flow cytometry plots and frequency of non-classical CD14^dim^CD16^+^ Mo as a proportion of live cells. Numbers indicate percentages in the drawn gates. (**G**) GeoMFI of CD38 in CD14^dim^CD16^+^ Mo. (**H**) Frequency of mDCs, (**I**) basophils, (**J**) pDCs, and (**K**) NK cells within total live cells. (**L**) GeoMFI of CD38 in NK cells. (**M**) Representative flow cytometry plots and frequency of CD56^bright^CD16^-^ NK cells. Numbers indicate percentages in the drawn gates. Data are presented as individual dots. Dark-colored plots show ICU patients. Bars represent mean ± SEM. Two-tailed Mann-Whitney U test was performed to compare HCs with COVT1 and HCs with COVT2. Wilcoxon matched pairs test was performed to compare COVT1 with COVT2 (*P < 0.05, **P < 0.01, and ***P < 0.001). HCs, *n*=21; COVT1, *n*=25; COVT2, *n*=20. (**D**,**E**,**G** and **L**) Data are presented in boxplots. Box boundaries represent the 1^st^ and 3^rd^ quartile of the distribution, while the center line represents the 2nd quartile (median). Whiskers go down to the smallest value and up to the largest.

We then analysed the frequency of CD14^+^CD16^-^ conventional monocytes, CD14^+^CD16^+^ intermediate monocytes and CD14^dim^CD16^+^ non-conventional monocytes (**Fig. 6A-C** and **Fig. S8A**). Classical CD14^+^CD16^-^ monocytes remained unaltered, whereas intermediate CD14^+^CD16^+^ monocytes showed a higher frequency in COVT2 compared to COVT1 and healthy controls (**Fig. S8D-E**). Furthermore, we observed a robust but transient decrease in the frequency of non-conventional CD14^dim^CD16^+^ monocytes (**Fig. 6B, C and F**), which appeared to be strongly activated during acute infection (**Fig. 6G**). Non-conventional monocytes have a patrolling function and contribute to the antiviral response^43^, which may point to their selective recruitment to the inflamed lung upon FcγRIIIA-mediated activation by virus-IgG immune complexes. Accordingly, non-conventional monocytes have been shown to be enriched in the lungs of critical COVID-19 patients^44^. The frequency of circulating myeloid dendritic cells (mDCs) and basophils were also strongly reduced in COVT1 compared to healthy controls, but both returned to homeostatic levels two months PSO (**Fig. 6H-I**). Interestingly, we detected increased CD25 expression on basophils from COVT1 compared to COVT2, which suggests basophil activation during the acute phase of infection (**Fig. S8F**).

Finally, we analyzed the frequency of circulating plasmacytoid DCs (pDCs) and natural killer (NK) cells, two subsets of the innate immune system that play crucial protective roles in viral infections. As reported previously^45^, the frequency of pDCs significantly decreased during acute infection (**Fig. 6J**). Remarkably, convalescent individuals maintained significantly lower frequency of pDCs compared to healthy controls, even after viral clearance (**Fig. 6J)**, suggesting persistent impaired type I IFN responses in infected individuals.

Although our data did not reveal any significant changes in the frequencies of natural killer (NK) cells within live cells, the analysis of CD38 expression indicated an activated phenotype during the acute phase of the infection (**Fig. 6K, L**). Moreover, the characterization of NK cell subsets based on the relative expression of CD56 and CD16 showed a persistent decrease in the frequency of circulating immature CD56^bright^CD16^-^ cells and an increase in the proportion of naturally cytotoxic CD56^dim^CD16^high^ cells within total NK cells (**Fig. 6M** and **Fig. S8G**). Thus, SARS-CoV-2 infection drives profound changes in the circulating innate immune compartment, some of which persist after viral clearance during convalescence.

### High dimensional analysis reveals coordinated immune responses and their association with disease severity

The immune system is composed of layered defense mechanisms of increasing specificity that protect the host from infection. To investigate the putative associations between innate and adaptive immune trajectories developed upon SARS-CoV-2 infection, we performed pairwise correlations across 41 variables that were significantly different among the groups analyzed, including virus-specific antibody titers measured by ELISA and selected immune parameters identified by high-dimensional flow cytometry (**Data File S1**). We found that the global antibody response to SARS-CoV-2 positively correlated with the frequency of circulating PCs, extrafollicular DN2 ME B cells, activated CD4^+^ T cells, cTfh cells and CD8^+^ T cells. It also positively correlated with CD38 expression on circulating myeloid cells, which suggests a close coordination between innate and adaptive immune responses. On the other hand, the level of virus-specific antibody titers, the percentage of circulating PCs and the frequency of CD19^+^ cells inversely correlated with the frequency of CD3^+^ T cells, non-conventional monocytes and pDCs, which suggests virus-induced interference with different components of both adaptive and innate immune responses in a fraction of COVID-19 patients (**Fig. 7A**). Next, principal component analysis (PCA) was performed to examine the general distribution of healthy controls and infected individuals during acute and convalescent stage according to these immune parameters. The analysis revealed that patients during acute infection were notably more distant from healthy controls than same patients during convalescence in PCA space (**Fig. 7B**). Virus-specific IgA titers, as well as the frequency of IgG and IgM PCs, activated CD4 T cells and pDCs, were among the variables that mostly contributed to the observed clustering in PC1 space. The frequency of naïve B cells and, to a lesser extent, IgM^+^IgD^+^ ME B cells, IgA1 PCs and NP-specific IgM titers were the immune features that mostly contributed to the clustering in PC2 space (**Fig. 7C, D**). Remarkably, when we computed the Euclidean distance from each COVID-19 patient at T1 to the centroid of healthy controls, we observed a positive correlation between Euclidean distance and disease severity defined according to the patient’s oxygen requirement and serum concentration of ferritin and lactate dehydrogenase (**Fig. 7E**). Of note, the same analysis performed on samples from COVT2 did not show equivalent correlation patterns (data not shown), which suggests that immunological recovery trajectories occur independently of disease severity status. Thus, cross-dataset correlation and principal component analysis reveals coordinated immune responses in COVID-19 patients as well as immune parameters associated with disease severity and inflammation.

**Fig. 7.**
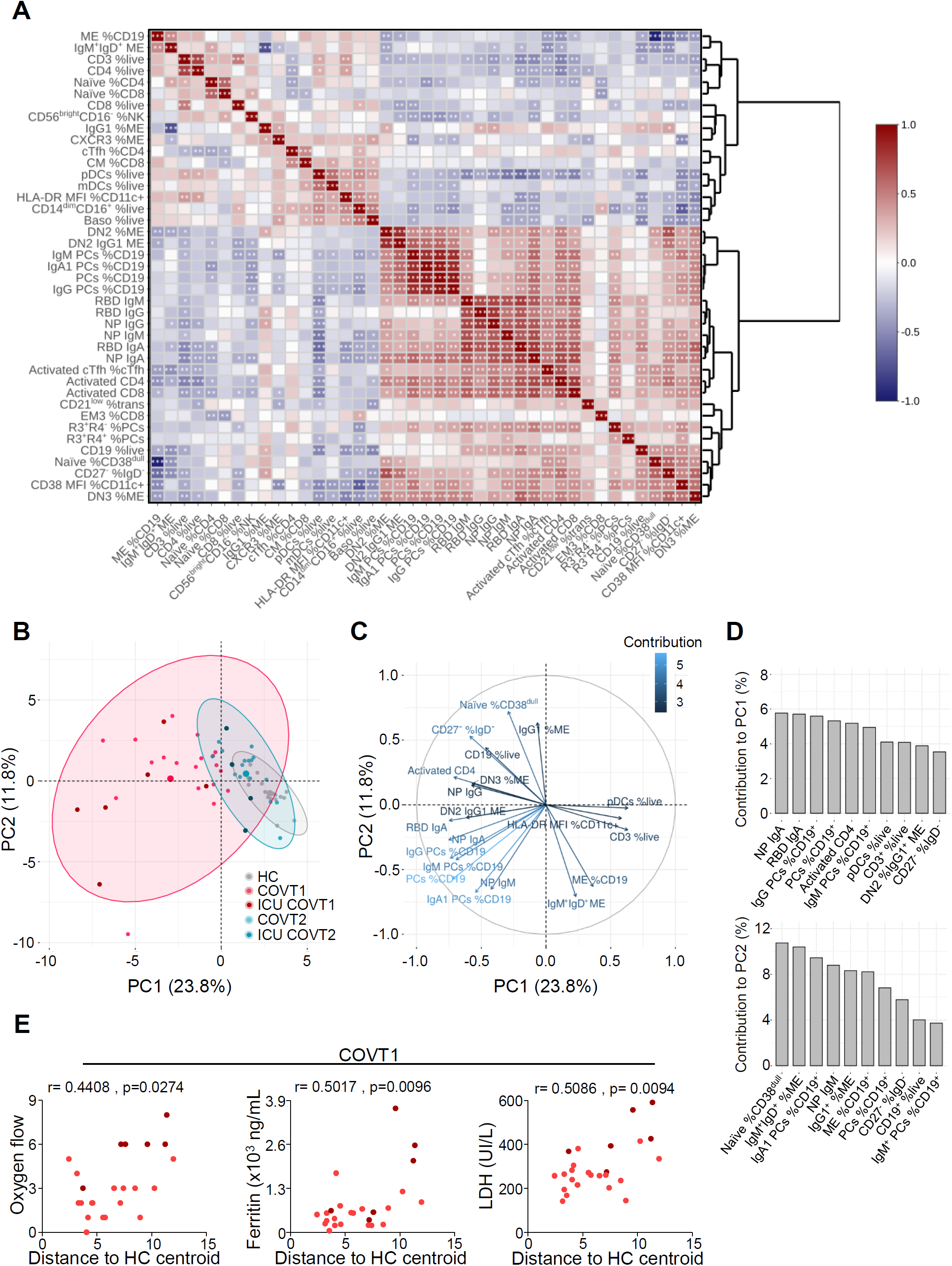
High-dimensional analysis of all variables studied reveals coordinated immune responses and their association with disease severity. (**A**) Spearman correlation mapping of indicated parameters for healthy controls (HCs), COVT1 and COVT2. Spearman’s rank correlation coefficient (ρ) was indicated by heat scale; Spearman p-value significance levels were corrected using Benjamini-Hochberg method significance (*P < 0.05, **P < 0.01, and ***P < 0.001). PCs, plasma cells. R3, CXCR3. R4, CXCR4. ME, memory. cTfh, circulating T follicular helper cells. CM, central memory. EM3, effector memory type 3. pDCs, plasmacytoid dendritic cells. mDCs, myeloid dendritic cells. Baso, basophils. NK, natural killer cells. trans, transitional B cells. (**B**) Principal component analysis (PCA) based on antibody titers, frequencies and GeoMFI of all measured markers (centered and scaled). RBD-specific parameters were excluded from the analysis. The first component explains 25% of the variation, whereas the second component explains 12.3% of the variation. Each dot represents a donor and each color represents a donor group. Dark-colored dots show ICU patients. Confidence ellipses for each group are plotted (confidence level set to 95%). (**C**) Variable correlation plot showing eigenvector-based coordinates of the top 20 variables in the two-dimensional space defined by the first two principal components. The relative position of the clinical variables reflects their relationship (positive correlated variables point to the same side of the plot; negative correlated variables point to opposite sides of the plot), while the length of the arrow is proportional to their contribution to the principal components. (**D**) Relative contribution of the top 10 variables to PC1 (top panel) and PC2 (bottom panel). (**A-D**) HCs, *n*=16; COVT1, *n*=25; COVT2, *n*=20. (**E**) Spearman correlation analysis of oxygen flow, ferritin and lactate dehydrogenase (LDH) levels in COVID-19 patients at COVT1 (*n*=25) plotted against the Euclidean distance from each COVT1 patient to the centroid of the HC group. The centroids by group were computed by averaging PC1 and PC2 for each member of group. The oxygen flow ranges from 0, meaning no need for supplemental oxygen, to 8, meaning the need for orotracheal intubation and mechanical ventilation. Data are presented as individual dots. Dark-colored dots show ICU patients. r stands for Spearman’s rank-order correlation.

## Discussion

A better understanding of immune changes occurring during COVID-19 progression and recovery is required to predict disease outcome and development of long-lasting immune protection. The vast majority of studies so far have focused on the immune profiling of elderly COVID-19 patients, who have an intrinsic risk to develop severe disease due to aging of the immune system and potential underlying health conditions. To circumvent this bias, we selected hospitalized patients younger than 65 and compared their immune parameters to equivalent parameters from age-matched healthy controls. This approach provided new insights into the breadth and kinetics of virus-specific B cell responses to SARS-CoV-2 and on the immunological landscape of not aged COVID-19 patients ranging from moderate to severe.

During acute SARS-CoV-2 infection, the presence of virus-specific antibodies with highly diversified Fc receptor-associated effector functions is consistent with an early involvement of multiple systemic and mucosal mechanisms of humoral protection, which presumably include immune pathways emerging from the gut^46^. The observed decline in RBD- and NP-specific IgM, IgA1 and IgA2 subclasses and the progressive increase in RBD-specific IgG1 during convalescence may reflect two temporal phases of humoral immunity to SARS-CoV-2: an initial stage characterized by a rapid release of poorly mutated and highly diversified antibody classes and subclasses, largely produced by short-lived PBs through extrafollicular pathways, followed by the emergence of high-affinity IgG generated by long-lived PCs. Indeed, the analysis of virus-specific B cell subset dynamics showed a transient increase in circulating RBD-specific HLA-DR^+^ PBs and extrafollicular DN B cells in acute infection, followed by a waning of early non-GC-derived B cell responses and sustained expansion of virus-specific IgG-dominated B cell memory. This breadth and kinetics of humoral responses may be set in place to consolidate robust antibody responses aimed at achieving complete viral clearance and long-lasting immune protection. Indeed, immune inhibitory signals generated by systemically produced monomeric IgA via the FcαRI receptor could interfere with the clearance of IgG-opsonized viral particles by phagocytes at a later phase of the infection. When engaged by monomeric forms of systemic IgA, the FcαRI receptor predominantly deploys inhibitory ITIM-mediated signals to phagocytes and NK cells^47^.

Another possible explanation for the reduction in virus-specific IgA1 and IgA2 two months PSO is the relocation of IgA-secreting PCs to mucosal inflamed tissues, including the upper respiratory tract. Indeed, acutely infected patients showed increased proportion of CXCR3^+^CXCR4^-^ PCs that are programmed to migrate to inflamed tissues rather than bone marrow niches. Consistently, two earlier reports described elevated SARS-CoV-2-specific IgA antibodies in nasal fluids, tears and saliva of infected individuals^27,48^. However, a recent study failed to detect PBs in the lung of deceased COVID-19 patients^49^.

Overall, our data are in agreement with recent reports showing detectable S-specific IgG and neutralizing antibodies at 6-8 months PSO and rapid decline of circulating IgA titers^14,15,17,18,33^. Moreover, we provided additional information on the dynamics of SARS-CoV-2-specific IgG subclasses in different phases of the infection. Compared to healthy controls, COVID-19 patients showed higher titers of all RBD- and NP-specific IgG subclasses, including poorly reported IgG2 and IgG4 subclasses, during acute infection and convalescence. Yet, only RBD-specific IgG1 increased over time, presumably because this antibody mostly originated from delayed germinal center responses. Finally, the magnitude and kinetics of IgG subclasses were highly heterogeneous between infected individuals. Thus, more studies with larger cohorts and later time points are required to explore the contribution of each IgG subclass to immune protection and recovery trajectories.

Recent reports have shown sustained development of virus-targeting ME B cells expressing IgG and a continuous evolution of the virus-specific antibody repertoire over time^14,17,33^, which indicates that SARS-CoV-2 infection promotes long-lasting GC-derived humoral responses. Similarly, we detected a sustained increase of RBD-specific IgG ME B cells that lasted for 6 months PSO. In addition, our analysis revealed that SARS-CoV-2 infection profoundly shaped the overall B cell memory compartment, promoting a sustained increase in the IgG1^+^ ME subset and a persistent contraction of unswitched CD27^+^IgM^+^IgD^+^ ME B cells. Loss of IgM^+^ ME B cells in COVID-19 patients has been previously described and associated with increased mortality and superimposed infections^50^. However, in our cohort of COVID-19 patients, the frequency of CD27^+^IgM^+^IgD^+^ ME B cells positively correlated with several markers of inflammation and NP-specific IgM titers. These results suggest that, in a subset of patients, humoral responses could diverge toward less efficient and non-neutralizing NP-targeting antibodies upon activation of B cell subsets previously primed by earlier seasonal coronavirus infections and therefore expressing cross-reactive but poorly protective IgM.

The mapping of global and SARS-CoV-2-specific B cell subsets in COVID-19 patients below 65 years of age also revealed some striking changes in mature B cell precursors. The increased frequency in CD21^low^ transitional and naïve B cells in COVT1 patients compared to age-matched healthy controls and COVT2 patients was consistent with the mobilization of B cell precursors to the periphery following virus-induced inflammation^51^. In the presence of massive immune sensing of viral particles, the immune system may be evolutionary programmed to recruit mature B cell precursors from the bone marrow into the periphery as a “last-ditch” defense against invading virions. Multiple inflammation-induced cytokines, including IL-7, may contribute to this process^34^.

In agreement with this hypothesis, we documented a significant expansion of RBD-specific naïve B cells enriched in Igλ that were largely activated and more numerous in COVID-19 patients with increased serum IL-7 and expanded naïve B cells. Altogether, these results support a scenario where *de novo* recruitment of mature B cell precursors to the periphery promotes the emergence of B cells expressing virus-reactive specificities constitutively engraved in the germline repertoire.

Consistent with this interpretation, stereotypic neutralizing antibodies with no or very few somatic mutations have been recently described in both the naïve and ME B cell repertoires^20–25^. Antibodies expressed by naïve and transitional B cells have notoriously different structural properties compared to mature antibodies expressed by GC-selected B cells. Compared to the latter, naïve and transitional B cells express antibodies that have a generally longer H-CDR3 in addition to increased autoreactivity and polyreactivity^52^. These properties may be central to the initial recognition and neutralization of the RBD-containing S protein on SARS-CoV-2. Antigen-activated naïve and transitional B cells could then transit through the GC to increase their affinity for viral antigens while redeeming their potential harmful autoreactivity. Therefore, the expansion of a naïve B cell repertoire expressing BCRs reactive to a wider range of neutralizing epitopes across the RBD of the S antigen may confer a functional advantage with respect to the initiation of protective antibody responses to SARS-CoV-2. However, the mobilization of B cell precursors to the periphery could also promote the emergence of pathogenic antibody responses to self-antigens due to defective central tolerance^51,52^. Indeed, the frequency of naïve B cells was a key variable to define distinct immune signatures that associated with disease severity and inflammation in COVID-19 patients. More studies are needed to evaluate longitudinal changes in the naïve Ig repertoire upon infection and their putative correlation with the onset of autoimmunity.

To further characterize the coordination of humoral immunity with other arms of the immune system in COVID-19 patients, we analyzed other circulating lymphoid and myeloid populations. Consistent with recent studies^12,53^, we showed robust activation of CD4^+^ and CD8^+^ T cells, including cTfh cells, in acutely infected patients and a strong correlation of T cell activation with several hallmarks of humoral responses. Our longitudinal analysis additionally revealed residual activation of the T cell pool as well as progressive expansion of cTfh, Th1, Th1/17 CD4^+^ T cells and subsets of memory CD8^+^ T cells during convalescence. These results suggest the continuous evolution of adaptive immunity to SARS-CoV-2 and are consistent with viral persistence in mucosal reservoirs or peripheral lymph nodes^18^.

In agreement with earlier reports (reviewed in ^54^), severe COVID-19 patients with increased inflammation also showed augmented T cell activation and humoral responses during acute infection. This observation may be explained by the presence of higher viral loads or prolonged periods of active viral replication in severe patients. However, a pathogenic role of adaptive immunity cannot be completely ruled out.

The role of innate immune responses in COVID-19 pathogenesis has been extensively described^42^. SARS-CoV-2 infection induces delayed and dysregulated IFN responses, thereby impairing proper priming of adaptive immune responses. In concordance with recent literature^45,55^, our data revealed a dramatic decrease of pDCs in acute infection that was marginally restored during convalescence. Although the depletion of circulating pDCs during acute infection could be suggestive of a targeted recruitment into virally infected tissues, their persistent lower frequency during convalescence argues in favor of a massive pDC apoptosis due to their activation through IFN signaling^56^. Moreover, being pDCs the main source of type I IFN, their loss may be linked to the aberrant interferon responses displayed by severe COVID-19 patients. Indeed, our study indicated that pDC depletion contributed to the definition of distinct immune signatures that correlated with poor prognosis and inflammation.

Moreover, as reported by others^5,57^, we found that acute SARS-CoV-2 infection markedly reduced HLA-DR expression on circulating myeloid cells and depleted non-conventional monocytes from circulation. Furthermore, our data revealed a negative correlation between HLA-DR expression and disease severity. Indeed, such decrease in HLA-DR expression has been recently associated to defective antigen-presentation driven by an excessive release of IL-6^5^. Finally, we found persistent changes in the circulating NK cell pool of patients infected with SARS-CoV-2. Our data are in line with previous reports suggesting a dysfunctional/exhausted phenotype of NK cells in COVID-19 patients as a consequence of hyper inflammation^58^.

In summary, our in-depth characterization of SARS-CoV-2-specific B cell responses revealed a previously unappreciated expansion of virus-specific naïve-like B cells over time, perhaps through the continuous mobilization of mature B cell precursors to the periphery. Moreover, our results consolidated previous findings on the immune response dynamics occurring in COVID-19 patients, showing both transient and long-lasting changes associated with disease severity and development of immune memory.

## Material and Methods

### Experimental Model and Subject Details Study Cohort

Blood samples were collected from COVID-19 patients (*n* = 25) in acute phase of infection (COVT1) hospitalized at the Hospital del Mar (Barcelona, Spain), with patient informed consent. Only patients with confirmed SARS-CoV-2 infection by reverse transcription quantitative polymerase chain reaction (RT-qPCR) of nasopharyngeal swab were included. All COVT1 samples were collected within the first 14 days following symptom onset, being the median time from symptom onset to blood testing 11 days [8-14]. A follow-up blood sample was collected from 20 out of the 25 initial participants two months after first sample collection (COVT2), being the median time from symptom onset to COVT2 blood testing 70 days [59-84]. The median age of patients was 51 years old. 52 % were males. The median length of stay from hospitalization to discharge was 16 days [5-90]. Clinical laboratory data were collected from the date closest to the first day of hospitalization. Blood samples from age-matched healthy donors (n = 21) were collected. All research subjects were pseudonymized by replacing directly identifiable variables with an ID immediately when collecting data. Patients ID were not known to anyone outside the research group. Demographic and clinical data of patients included in the study are summarized in **Table S1**. All procedures followed were approved by the Ethical Committee for Clinical Investigation of the Institut Hospital del Mar d’Investigacions Mèdiques (Number 2020/9189/I).

### Sample collection and processing

For all COVID-19 patients and healthy volunteers, sera were collected from whole blood in silica-treated tubes (BD Biosciences) where the blood was incubated for 30 min without movement to trigger coagulation. Next, samples were centrifuged for 10 min at 1300 g at room temperature (RT), heat-inactivated at 56°C for 1 hour and stored at −80°C prior to use. PBMCs were isolated from whole blood collected with EDTA anticoagulant via Ficoll-Paque Premium (Cytiva) following manufacturer’s instructions. Briefly, each sample was diluted with an equal volume of phosphate buffered saline (PBS). The diluted blood was carefully layered over an equal volume of Ficoll solution in a Falcon tube, followed by centrifugation at 400 g for 30 min at 20°C without brake. The upper layer containing plasma was collected and stored at −80°C. The layer of mononuclear cells was gently removed and washed twice with PBS. Pelleted cells were counted using Turk solution, resuspended in fetal bovine serum (FBS, Gibco) with 10% Dimethyl sulfoxide (DMSO, Sigma) and stored in liquid nitrogen prior to use.

### Production of recombinant SARS-CoV-2 proteins

The pCAGGS RBD construct, encoding for the receptor-binding domain of the SARS-CoV-2 Spike protein (amino acids 319-541 of the Spike protein) along with the signal peptide plus a hexahistidine tag was provided by Dr Krammer (Mount Sinai School of Medicine, NY USA). The pLVX-EF1alpha-nCoV2019-N-2xStrep-IRES-Puro construct, encoding for the full-length SARS-CoV-2 nucleocapsid protein (NP) fused to a double Strep-tag at the C-terminus was a gift from Dr Krogan (University of California, San Francisco USA). Recombinant proteins were expressed in-house in Expi293F human cells (Thermo Fisher Scientific) by transfection of the cells with purified DNA and polyethylenimine (PEI). For secreted RBD proteins, cells were harvested 3 days post transfection and RBD-containing supernatants were collected by centrifugation at 13000rpm for 15min. RBD proteins were purified in Hitrap-ni Columns in an automated Fast Protein Liquid Chromatography (FPLC; Äkta avant), concentrated through 10 kDa Amicon centrifugal filter units (EMD Millipore) and resuspended in phosphate buffered saline (PBS). For NP, cell lysates from transfected cells were centrifuged at 13000 rpm for 20 min and the supernatant was loaded into a 5 ml Strep-Tactin XT column. The eluted nucleocapsid was concentrated through a 10 kDa Amicon centrifugal filter unit and purified with SEC (Sephadex 10/300) in PBS.

### Enzyme-linked immunosorbent assay (ELISA)

ELISAs performed in this study were adapted from previously established protocols^46^. Serum samples were heat-inactivated at 56°C for 1 hour and stored at −80°C prior to use. 96-well half-area flat bottom high-bind microplates (Corning) were coated overnight at 4°C with each respective recombinant viral protein at 2µg/ml in PBS (30 µl per well) or with PBS alone. Plates were washed with PBS 0.05% Tween 20 (PBS-T) and blocked with blocking buffer (PBS containing 1.5% Bovine serum albumin, BSA) for 2 hours at RT. Serum samples were serially diluted in PBS supplemented with 0.05% Tween 20 and 1% BSA and added to the viral protein- or PBS-coated plates for 2 hours at RT. After washing, plates were incubated with horseradish peroxidase (HRP)-conjugated anti-human Ig secondary antibodies diluted in PBS containing 0.05% Tween 20 1% BSA for 45 minutes at RT. Plates were washed 5 times with PBS-T and developed with TMB substrate reagent set (BD bioscience) with development reaction stopped with 1M H2SO4. Absorbance was measured at 450nm on a microplate reader (Infinite 200 PRO, Tecan). To detect RBD-specific and NP-specific IgM, IgA and IgG, HRP-conjugated anti-human IgA, IgG and IgM (Southern Biotech) were used at a 1:4000 dilution. To assess the distribution of the different IgG antibody subclasses, HRP-conjugated anti-human IgG1, IgG2, IgG3 and IgG4 (Southern Biotech) were used at a 1:3000 dilution. To detect SARS-CoV-2-specific IgA1 and IgA2, HRP-conjugated anti-human IgA1 or IgA2 (Southern Biotech) were used at a dilution of 1:2000 and 1:4000, respectively. To quantitate the level of each viral antigen-specific antibody class or subclasses optical density (OD) values were calculated after subtraction of background (OD450 of serum dilutions on PBS-coated plates) and the area under the curve (AUC) derived from optical density measurements of six serial dilutions was determined using Prism 8 (GraphPad). AUC values below an established cutoff were replaced by the AUC value of the cutoff for plotting and calculation purposes. Negative threshold values were set using healthy control AUC levels plus 2 times the standard deviations of the mean.

The concentration of interleukin-7 (IL-7) was measured by a sandwich ELISA kit (Peprotech) following the manufacturer’s instructions on 50 µl plasma samples. The levels of IL-7 were expressed as pg/mL of plasma.

### Spectral flow cytometry

For spectral flow cytometry, frozen Peripheral Blood Mononuclear Cells (PBMCs) were thawed in warm complete RPMI 1640 medium (Biowest) and centrifuged at 1450 rpm for 5 min at RT. Cells were then resuspended in 2 mL of LIVE/DEAD Fixable Yellow Dead Cell Stain Kit (Thermo Fisher Scientific) 1:20000 in PBS, incubated for 30 min at RT and stained with two different fluorophore-conjugated antibody cocktails (**Table S2** and S**4**). Only for MIX 3, cells were washed and resuspended in 50 µL cool staining buffer with Fc-receptor blocking reagent for 10 min at 4 °C. For staining, a sequential approach was followed. Anti-CCR7 antibody was added first and incubated for 10 min. Then, all other anti-chemokine receptor antibodies (CXCR3, CXCR4, CXCR5, CCR4 and CCR6 for MIX3, and CXCR3 and CXCR4 for MIX 1) were added to the corresponding tubes and incubated for 10 min. After that, the remaining antibodies were added and incubated for 20 min (MIX 1) or 30 min (MIX 3). Stained cells were washed and resuspended in 4% paraformaldehyde in PBS while vortexing, and incubated for 12 min in ice. Fixed cells were then washed, resuspended in PBS and acquired using the Aurora spectral analyzer (Cytek). Data was analyzed using FlowJo V10.6.2 software (TreeStar).

### Flow cytometric detection of SARS-CoV-2-RBD-specific B cells

1.5 million frozen PBMCs were thawed, centrifuged and resuspended in a 1:20000 PBS dilution of LIVE/DEAD Fixable Yellow Dead Cell Stain Kit (Thermo Fisher Scientific) for 30 min at RT to exclude dead cells. Cells were then washed with PBS and incubated with 13.2 pmol SARS-CoV-2 RBD-HIS Biotinylated Recombinant Protein (Sino Biological) diluted in 50 µl 1X PBS supplemented with 0.2% BSA and 2 mM EDTA for 30 min in ice. Cells were washed again and stained with the MIX 2 antibody cocktail (**Table S3**) plus 1.32 pmol Streptavidin Alexa Fluor 647 (Thermo Fisher Scientific) for 30 min. Stained cells were washed, fixed with 4% paraformaldehyde in PBS for 12 min and acquired with LSR Fortessa (BD Biosciences). Data were further analyzed with FlowJo V10.6.2 software.

Alternatively, 6.6 pmol SARS-CoV-2 RBD-HIS Biotinylated Recombinant Protein was incubated for 1 h in PBS with 0.94 pmol Streptavidin Alexa Fluor 647 and 0.94 pmol Streptavidin Alexa Fluor 488 (Thermo Fisher Scientific), separately. Meanwhile, 1.5 million frozen PBMCs were thawed, centrifuged and resuspended in 50 µl PBS. Next, cells were incubated in the same staining tube with the labeled RBD probes for 20 min in ice. Cells were then washed and stained with the MIX 2 antibody cocktail for 10 min using anti-human IgA AmCyan instead of anti-human IgA FITC (**Table S3**) and DAPI fluorescent dye (Sigma Aldrich). Stained cells were washed and resuspended in PBS 2%FBS and acquired on LSR Fortessa (BD Bioscience).

### High-dimensional data analysis of flow cytometry data

t-Distributed Stochastic Neighbor Embedding (tSNE) analyses were performed with Flowjo V10.6.2 software. tSNE analysis was performed using equal sampling from each FCS file, with 1000 iterations, a perplexity of 30, a Barnes-Hut gradient algorithm and exact KNN algorithm. For CD19^+^ cells, the following samples were concatenated: COVT1/T2 (*n* = 16) and healthy controls (*n* = 11); and the following markers from MIX 1 were used to generate the tSNE maps: CXCR4, CXCR3, CD10, CD19, IgM, CD27, CD21, IgA1+IgA2, IgD, HLA-DR, CD38, CD24, CD43, CD11c, CD45RB and LIVE/DEAD YELLOW. For both CD19^-^CD3^+^ and CD19^-^CD3^-^ cells, all samples were included in the analysis and the following markers from MIX 2 were used: CCR7, CXCR5, CCR6, CXCR3, CCR4, CD16, CD3, CD8a, CD11c, CD123, CD56, PD-1, CD14, CD45RA, HLA-DR, ICOS, CD4, CD25, CD19, CD27, CCR4, CD127, CD38 and LIVE/DEAD YELLOW.

### Data analysis and visualization

GraphPad Prism (version 8.0) and R (version 3.6.3, R Core Team (2019). R: A Language and Environment for Statistical Computing) were used to conduct statistical analyses. For each experiment, the type of statistical test, summary statistics and levels of significance were specified in the figures and corresponding legends. The correlation analysis was performed in R. We performed data imputation of the missing variables using predictive mean matching implemented in the mice package^59^. Pairwise Spearman’s rank correlations between 41 variables (including antibody titers measured by ELISA and immune parameters defined by high-dimensional flow cytometry; **Data File S1**) for COVT1 (*n* = 25), COVT2 (*n* = 20) and healthy controls (*n* = 16) were calculated and visualized as a correlogram using R package corrplot (Taiyun Wei and Viliam Simko (2017). R package “corrplot": Visualization of a Correlation Matrix). Spearman’s rank correlation coefficient (ρ) was indicated by heat scale; significance was indicated by *P < 0.05, **P < 0.01, and ***P < 0.001; FDR correction was performed using the Benjamini-Hochberg procedure at the FDR < 0.05 significance threshold.

Principal component analysis (PCA) was used to identify the most important features from 41 variables (including antibody titers and immune parameters; **Data File S1**) using COVT1 (*n* = 25), COVT2 (*n* = 20) and healthy controls (*n* = 16). The PCA was conducted using the “prcomp” function in R and visualized using the “factoextra” package (Kassambara A, and Mundt F. factoextra: Extract and Visualize the Results of Multivariate Data Analyses. 2020).

## Supporting information

de Campos-Mata supplementary materials

## Data Availability

All data are available in the main text or the supplementary materials. Further information and requests for resources and reagents should be directed to and will be fulfilled by the corresponding author, Giuliana Magri.

## Acknowledgments

We want to particularly acknowledge the patients and the Parc de Salut Mar MARBiobanc (PT17/0015/0011) integrated in the Spanish National Biobanks Network from ISCIII for their collaboration. MARBiobanc’s work was supported by grants from Instituto de Salud Carlos III/FEDER (PT17/0015/0011) and the “Xarxa de Bancs de tumors” sponsored by Pla Director d’Oncologia de Catalunya (XBTC). This study was supported by the COVID-19 call grant from Generalitat de Catalunya, Department of Health (to G.M and L.D.C.M) and grant Miguel Servet research program (to G.M). IMI-JU resources of which are composed of financial contribution from the EU-FP7 [FP7/2007–2013] and EFPIA companies in kind contribution [116030 to TransQST, 777365 to eTRANSAFE], and the EU H2020 Programme 2014–2020 [676559 to Elixir-Excelerate]; Agència de Gestió d’Ajuts Universitaris i de Recerca Generalitat de Catalunya [2017SGR00519]. The Research Programme on Biomedical Informatics (GRIB) is a member of the Spanish National Bioinformatics Institute (INB), funded by ISCIII and FEDER (PRB2-ISCIII [PT13/0001/0023, of the PE I+D+i 2013–2016]). The DCEXS is a ‘Unidad de Excelencia María de Maeztu’, funded by the MINECO [MDM-2014-0370].

## Author Contributions

L.D.C.M. designed and performed experiments, analyzed and discussed data and wrote the manuscript; J.P. performed data analysis; S.T.V. and R.T.P. designed and performed experiments and discussed data; N.R.M. and C.C. produced recombinant SARS-CoV-2 antigens; E.K.G. and A.C. discussed data and wrote the manuscript; J.V.G., J.P.H. and I.A.A. selected COVID-19 patients, provided clinical data and discussed data; G.M. designed and performed experiments, analyzed results, discussed data, and wrote the manuscript.

## References

1. WHO Coronavirus Disease (COVID-19) Dashboard. https://covid19.who.int/.

2. Guan, W. et al. Clinical Characteristics of Coronavirus Disease 2019 in China. N. Engl. J. Med. 382, 1708–1720 (2020).

3. Mehta, P. et al. COVID-19: consider cytokine storm syndromes and immunosuppression. The Lancet vol. 395 1033–1034 (2020).

4. Tay, M. Z., Poh, C. M., Rénia, L., MacAry, P. A. & Ng, L. F. P. The trinity of COVID-19: immunity, inflammation and intervention. Nature Reviews Immunology vol. 20 363–374 (2020).

5. Giamarellos-Bourboulis, E. J. et al. Complex Immune Dysregulation in COVID-19 Patients with Severe Respiratory Failure. Cell Host Microbe 27, 992-1000.e3 (2020).

6. Hadjadj, J. et al. Impaired type I interferon activity and inflammatory responses in severe COVID-19 patients. Science (80-.). 369, 718–724 (2020).

7. Mangalmurti, N. & Hunter, C. A. Cytokine Storms: Understanding COVID-19. Immunity vol. 53 19–25 (2020).

8. Bastard, P. et al. Autoantibodies against type I IFNs in patients with life-threatening COVID-19. Science (80-.). 370, (2020).

9. Zuo, Y. et al. Prothrombotic autoantibodies in serum from patients hospitalized with COVID-19. Sci. Transl. Med. 12, (2020).

10. Wang, E. Y. et al. Diverse functional autoantibodies in patients with COVID-19. medRxiv 2020.12.10.20247205 (2020).

11. Le Bert, N. et al. SARS-CoV-2-specific T cell immunity in cases of COVID-19 and SARS, and uninfected controls. Nature 584, 457–462 (2020).

12. Mathew, D. et al. Deep immune profiling of COVID-19 patients reveals distinct immunotypes with therapeutic implications. Science (80-.). 369, (2020).

13. Peng, Y. et al. Broad and strong memory CD4+ and CD8+ T cells induced by SARS-CoV-2 in UK convalescent individuals following COVID-19. Nat. Immunol. 21, 1336–1345 (2020).

14. Dan, J. M. et al. Immunological memory to SARS-CoV-2 assessed for up to 8 months after infection. Science (80-). eabf4063 (2021).

15. Wajnberg, A. et al. Robust neutralizing antibodies to SARS-CoV-2 infection persist for months. Science (80-.). 370, 1227–1230 (2020).

16. Rodda, L. B. et al. Functional SARS-CoV-2-Specific Immune Memory Persists after Mild COVID-19. Cell 184, 169-183.e17 (2021).

17. Hartley, G. E. et al. Rapid generation of durable B cell memory to SARS-CoV-2 spike and nucleocapsid proteins in COVID-19 and convalescence. Sci. Immunol. 5, (2020).

18. Gaebler, C. et al. Evolution of antibody immunity to SARS-CoV-2. Nature 1–6 (2021).

19. Sokal, A. et al. Maturation and persistence of the anti-SARS-CoV-2 memory B cell response. Cell (2021).

20. Kreer, C. et al. Longitudinal Isolation of Potent Near-Germline SARS-CoV-2-Neutralizing Antibodies from COVID-19 Patients. Cell vol. 182 1663–1673 (2020).

21. Kim, S. Il et al. Stereotypic neutralizing VHantibodies against SARS-CoV-2 spike protein receptor binding domain in patients with COVID-19 and healthy individuals. Sci. Transl. Med. 13, (2021).

22. Robbiani, D. F. et al. Convergent antibody responses to SARS-CoV-2 in convalescent individuals. Nature 584, 437–442 (2020).

23. Barnes, C. O. et al. Structures of Human Antibodies Bound to SARS-CoV-2 Spike Reveal Common Epitopes and Recurrent Features of Antibodies. Cell 182, 828-842.e16 (2020).

24. Seydoux, E. et al. Analysis of a SARS-CoV-2-Infected Individual Reveals Development of Potent Neutralizing Antibodies with Limited Somatic Mutation. Immunity (2020).

25. Schultheiß, C. et al. Next-Generation Sequencing of T and B Cell Receptor Repertoires from COVID-19 Patients Showed Signatures Associated with Severity of Disease. Immunity 53, 442-455.e4 (2020).

26. Feldman, J. et al. Naive human B cells can neutralize SARS-CoV-2 through recognition of its receptor binding domain. bioRxiv Prepr. Serv. Biol. (2021).

27. Sterlin, D. et al. IgA dominates the early neutralizing antibody response to SARS-CoV-2. Sci. Transl. Med. eabd2223 (2020).

28. Woodruff, M. C. et al. Extrafollicular B cell responses correlate with neutralizing antibodies and morbidity in COVID-19. Nat. Immunol. 21, 1506–1516 (2020).

29. Sanz, I. et al. Challenges and opportunities for consistent classification of human b cell and plasma cell populations. Frontiers in Immunology vol. 10 2458 (2019).

30. Muehlinghaus, G. et al. Regulation of CXCR3 and CXCR4 expression during terminal differentiation of memory B cells into plasma cells. Blood 105, 3965–3971 (2005).

31. Cain, D., Kondo, M., Chen, H. & Kelsoe, G. Effects of acute and chronic inflammation on B-cell development and differentiation. Journal of Investigative Dermatology vol. 129 266–277 (2009).

32. Groom, J. R. & Luster, A. D. CXCR3 ligands: Redundant, collaborative and antagonistic functions. Immunology and Cell Biology vol. 89 207–215 (2011).

33. Rodda, L. B. et al. Functional SARS-CoV-2-Specific Immune Memory Persists after Mild COVID-19. Cell 184, 169-183.e17 (2021).

34. Ceredig, R. & Rolink, A. G. The key role of IL-7 in lymphopoiesis. Seminars in Immunology vol. 24 159–164 (2012).

35. Chailyan, A., Marcatili, P., Cirillo, D. & Tramontano, A. Structural repertoire of immunoglobulin λ light chains. Proteins Struct. Funct. Bioinforma. 79, 1513–1524 (2011).

36. Chen, Z. & John Wherry, E. T cell responses in patients with COVID-19. Nat. Rev. Immunol. 20, 529–536 (2020).

37. Wilkinson, T. M. et al. Preexisting influenza-specific CD4 + T cells correlate with disease protection against influenza challenge in humans. Nat. Med. 18, 274–280 (2012).

38. Miller, J. D. et al. Human Effector and Memory CD8+ T Cell Responses to Smallpox and Yellow Fever Vaccines. Immunity 28, 710–722 (2008).

39. Crotty, S. T Follicular Helper Cell Biology: A Decade of Discovery and Diseases. Immunity vol. 50 1132–1148 (2019).

40. Morita, R. et al. Human Blood CXCR5+CD4+ T Cells Are Counterparts of T Follicular Cells and Contain Specific Subsets that Differentially Support Antibody Secretion. Immunity 34, 108–121 (2011).

41. Vella, L. A. et al. T follicular helper cells in human efferent lymph retain lymphoid characteristics. J. Clin. Invest. 129, 3185–3200 (2019).

42. Schultze, J. L. & Aschenbrenner, A. C. COVID-19 and the human innate immune system. Cell (2021).

43. Narasimhan, P. B., Marcovecchio, P., Hamers, A. A. J. & Hedrick, C. C. Nonclassical Monocytes in Health and Disease. Annual Review of Immunology vol. 37 439–456 (2019).

44. Sánchez-Cerrillo, I. et al. COVID-19 severity associates with pulmonary redistribution of CD1c+ DCs and inflammatory transitional and nonclassical monocytes. J. Clin. Invest. 130, 6290–6300 (2020).

45. Zhou, R. et al. Acute SARS-CoV-2 Infection Impairs Dendritic Cell and T Cell Responses. Immunity 53, 864-877.e5 (2020).

46. de Campos Mata, L. et al. SARS-CoV-2-specific antibody profiles distinguish patients with moderate from severe COVID-19. medRxiv 2020.12.18.20248461 (2020).

47. Pasquier, B. et al. Identification of FcαRI as an inhibitory receptor that controls inflammation: Dual role of FcRγ ITAM. Immunity 22, 31–42 (2005).

48. Cervia, C. et al. Systemic and mucosal antibody responses specific to SARS-CoV-2 during mild versus severe COVID-19. J. Allergy Clin. Immunol. 147, 545-557.e9 (2021).

49. Ferreira-Gomes, M. et al. SARS-CoV-2 in severe COVID-19 induces a TGF-β-dominated chronic immune response that does not target itself. Nat. Commun. 12, 1961 (2021).

50. Lenti, M. V. et al. Depletion of circulating IgM memory B cells predicts unfavourable outcome in COVID-19. Sci. Rep. 10, (2020).

51. Cain, D., Kondo, M., Chen, H. & Kelsoe, G. Effects of acute and chronic inflammation on B-cell development and differentiation. Journal of Investigative Dermatology vol. 129 266–277 (2009).

52. Wardemann, H. et al. Predominant autoantibody production by early human B cell precursors. Science (80-.). 301, 1374–1377 (2003).

53. Weiskopf, D. et al. Phenotype and kinetics of SARS-CoV-2-specific T cells in COVID-19 patients with acute respiratory distress syndrome. Sci. Immunol. 5, (2020).

54. Brodin, P. Immune determinants of COVID-19 disease presentation and severity. Nature Medicine vol. 27 28–33 (2021).

55. Kuri-Cervantes, L. et al. Comprehensive mapping of immune perturbations associated with severe COVID-19. Sci. Immunol. 5, (2020).

56. Swiecki, M. et al. Type I interferon negatively controls plasmacytoid dendritic cell numbers in vivo. J. Exp. Med. 208, 2367–2374 (2011).

57. Schulte-Schrepping, J. et al. Severe COVID-19 Is Marked by a Dysregulated Myeloid Cell Compartment. Cell 182, 1419-1440.e23 (2020).

58. van Eeden, C., Khan, L., Osman, M. S. & Tervaert, J. W. C. Natural killer cell dysfunction and its role in covid-19. International Journal of Molecular Sciences vol. 21 1–17 (2020).

59. van Buuren, S. & Groothuis-Oudshoorn, K. mice: Multivariate imputation by chained equations in R. J. Stat. Softw. 45, 1–67 (2011).

