## Supplementary material for "SARS-CoV-2 sculpts the immune system to induce sustained virus-specific naïve-like and memory B cell responses": de Campos-Mata supplementary materials

Fig. S1 (related to Fig.1). SARS-CoV-2 infection induces RBD- and NP-specific humoral responses.

Fig. S2 (related to Fig. 2). Additional characterization of plasma cell and naïve B cell phenotype in COVID-19 patients.

Fig. S3 (related to Fig. 3). SARS-CoV-2 drives an expansion of unconventional $IgD^-$ $CD27^-$ DN B cells in the acute phase of infection.

Fig. S4 (related to Fig. 4). SARS-CoV-2 infection induces sustained RBD-specific $IgG^+$ memory B cells.

Fig. S5 (related to Fig. 4). Long-term analysis of COVID-19 convalescent individuals suggest persistent expansion of RBD-specific $IgG^+$ memory B cells up to six months post-infection.

Fig. S6 (related to Fig. 5). Gating strategy for major T cell populations.

Fig. S7 (related to Fig. 5). SARS-CoV-2 infection associates with changes in the relative frequency of $CD4^+$ T helper subsets.

Fig. S8 (related to Fig. 6). SARS-CoV-2 infection induces long-lasting changes in the innate immune compartment.

Table S1. Demographic and clinical data of patients included in the study.

Table S2. Antibodies used for phenotypical characterization of B cell subsets by high-dimensional flow cytometry.

Table S3. Antibodies used for flow cytometric detection and phenotypical characterization of SARS-CoV-2-RBD-specific B cells.

Table S4. Antibodies used for phenotypical characterization of other circulating lymphoid and myeloid populations by high-dimensional flow cytometry.

Data file S1. Raw data of SARS-CoV-2-specific antibody titers and selected immune parameters used to perform pairwise correlations and PCA (provided as a separate Excel file).

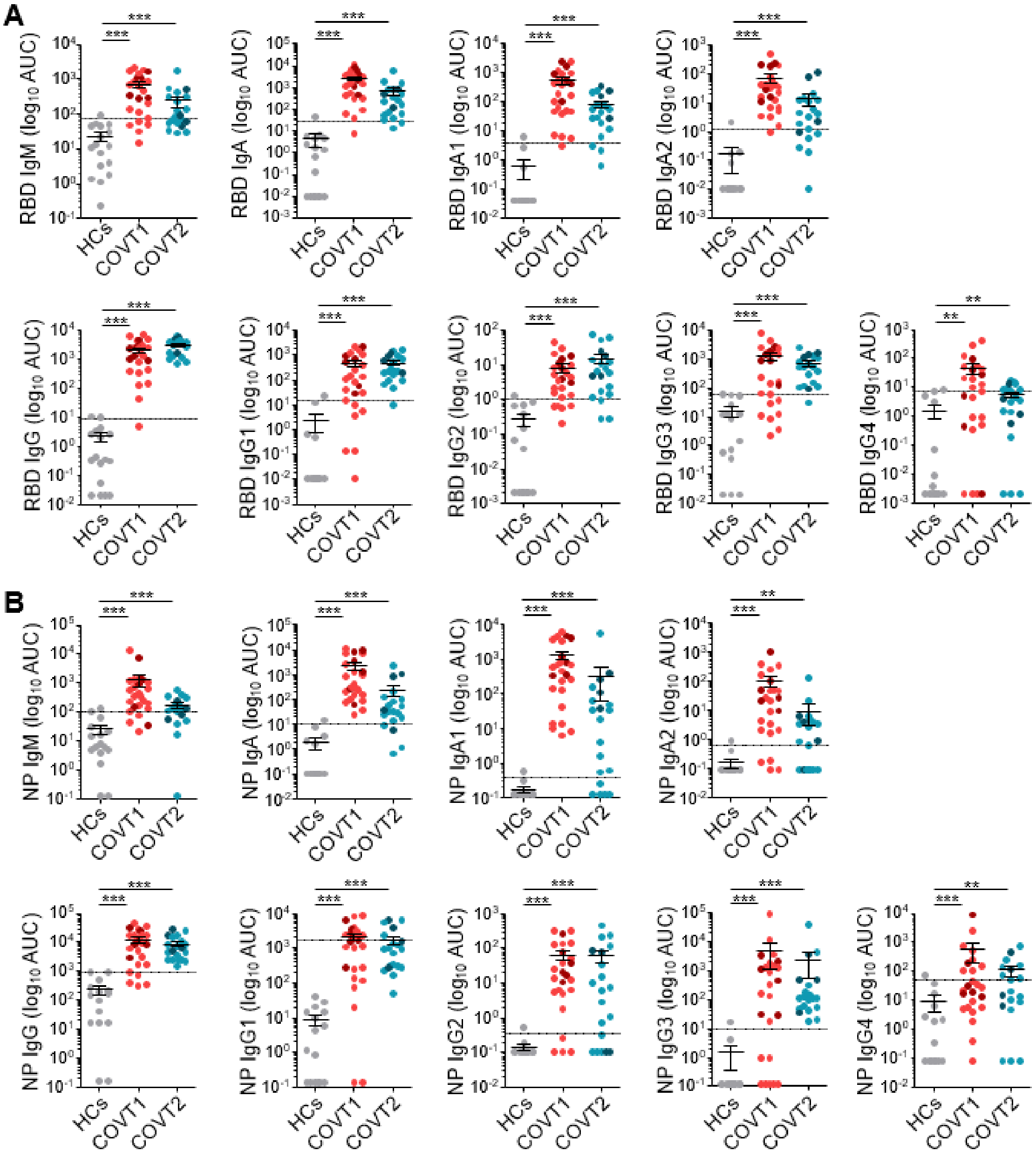


**Fig. S1 (related to Fig. 1). SARS-CoV-2 infection induces RBD- and NP-specific humoral responses.** (**A**) Area under the curve (AUC) for each of the RBD-specific and (**B**) NP-specific antibody classes and subclasses in sera samples from healthy controls (HCs) and COVID-19 patients in the acute (COVT1) and convalescent phase (COVT2). Sera from HCs were analyzed to establish negative threshold values defined as the HC AUC mean plus 2 times the standard deviation of the HC mean. Dashed line indicates negative threshold. Data are presented as individual dots. Dark-colored dots show ICU patients. Bars represent mean ± SEM. Two-tailed Mann-Whitney U test was performed to compare HCs with COVT1 and HCs with COVT2 (*P < 0.05, **P < 0.01, and ***P < 0.001). HCs, *n*=17; COVT1, *n*=25; COVT2, *n*=20.

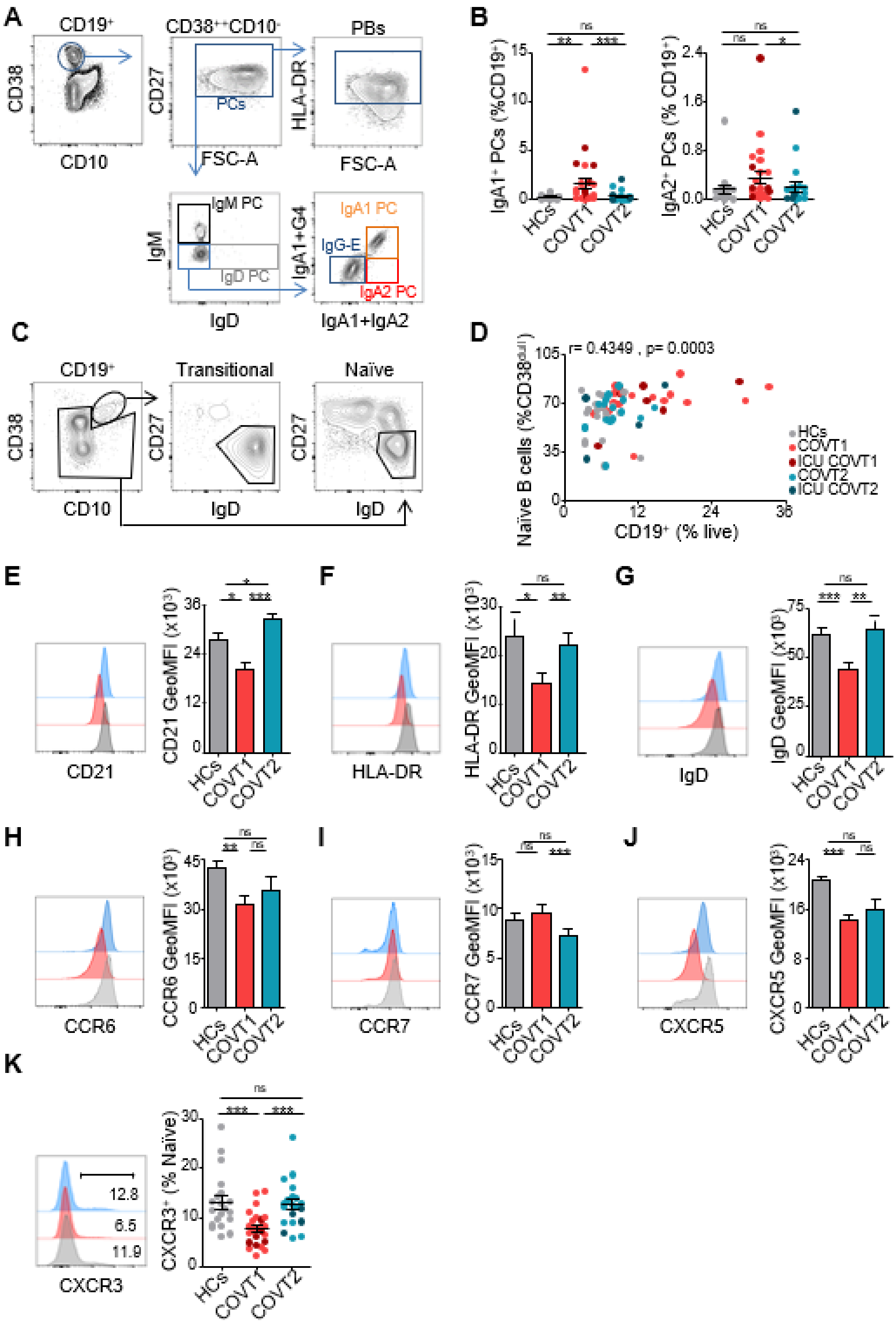

**Fig. S2 (related to Fig. 2). Additional characterization of plasma cell and naïve B cell phenotype in COVID-19 patients.** (**A**) Gating strategy used to define circulating plasma cell (PC) subsets. (**B**) Frequencies of PCs expressing IgA1 and IgA2 isotypes from total CD19$^{+}$ B cells. (**C**) Gating strategy used to define transitional (CD38$^{int}$CD10$^{+}$IgD$^{+}$CD27$^{-}$) and naïve (IgD$^{+}$CD27$^{-}$) B cells. (**D**) Spearman correlation analysis of naïve B cells within CD19$^{+}$CD38$^{dull}$ cells and CD19$^{+}$ cells within live lymphocytes in all samples analyzed. r stands for Spearman's rank-order correlation. (**E**) Representative flow cytometry histogram and Geometric Mean Fluorescence Intensity (GeoMFI) of CD21, (**F**) HLA-DR and (**G**) IgD expression in $IgD^{+}CD27^{-}$ naïve B cells. (**H**) Representative flow cytometry histogram and GeoMFI of CCR6, (**I**) CCR7 and (**J**) CXCR5 expression in $CD27^{-}$ B cells. (**K**) Representative flow cytometry histogram and frequency of naïve B cells expressing CXCR3. Data are presented as mean ± SEM. Two-tailed Mann-Whitney U test was performed to compare healthy controls (HCs) with COVT1 and HCs with COVT2. Wilcoxon matched pairs test was performed to compare COVT1 with COVT2 (*P < 0.05, **P < 0.01, and ***P < 0.001). HCs, *n*=19; COVT1, *n*=25; COVT2, *n*=20. (**B,D,K**) Data are presented as individual dots. Dark-colored dots show ICU patients. Bars represent mean ± SEM.

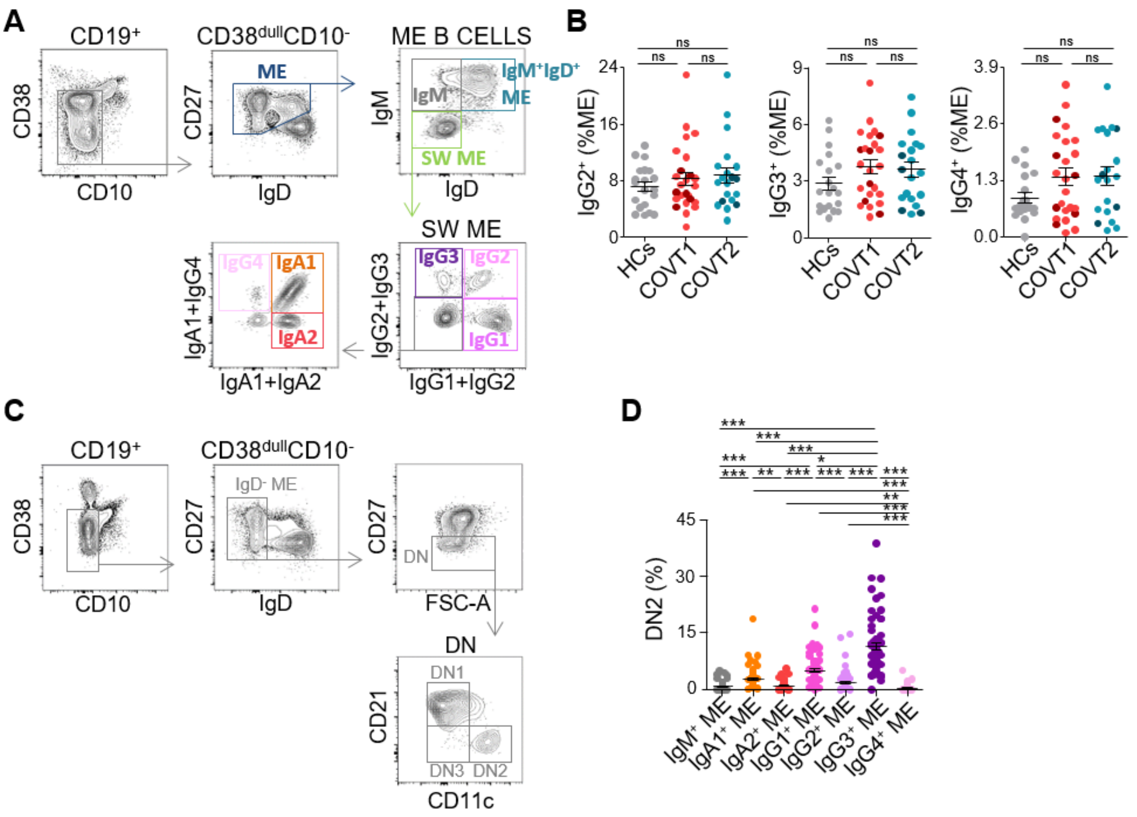


**Fig. S3 (related to Fig. 3). SARS-CoV-2 drives an expansion of unconventional $IgD^{-}$ $CD27^{-}$ DN B cells in the acute phase of infection.** (**A**) Gating strategy used to define the following memory (ME) B cell populations: ME ($CD38^{dull}CD10^{-}IgD^{-}$), $IgM^{+}$ ME ($CD38^{dull}CD10^{-}IgD^{-}IgM^{+}$), $IgM^{+}IgD^{+}$ ME ($CD38^{dull}CD10^{-}IgD^{+}IgM^{+}$), switched (SW) ME ($CD38^{dull}CD10^{-}IgD^{-}IgM^{-}$) and subtypes. (**B**) Frequency of $IgG2^{+}$, $IgG3^{+}$ and $IgG4^{+}$ ME B cells within total ME B cells in healthy controls (HCs), COVT1 and COVT2. (**C**) Gating strategy used to define DN1 ($IgD^{-}CD27^{-}CD21^{+}CD11c^{-}$), DN2 ($IgD^{-}CD27^{-}CD21^{-}CD11c^{+}$) and DN3 ($IgD^{-}CD27^{-}CD21^{-}CD11c^{-}$) B cell populations. (**D**) Frequencies of DN2 cells among different ME B cell subsets in HCs, COVT1 and COVT2 merged samples. Data are presented as individual dots. Dark-colored dots show ICU patients. Bars represent mean ± SEM. Two-tailed Mann-Whitney U test was performed to compare HCs with COVT1 and HCs with COVT2. Wilcoxon matched pairs test was performed to compare COVT1 with COVT2. (**D**) Kruskal-Wallis *H* test (*$P < 0.05$, **$P < 0.01$, and ***$P < 0.001$). HCs, *n*=19; COVT1, *n*=25; COVT2, *n*=20.

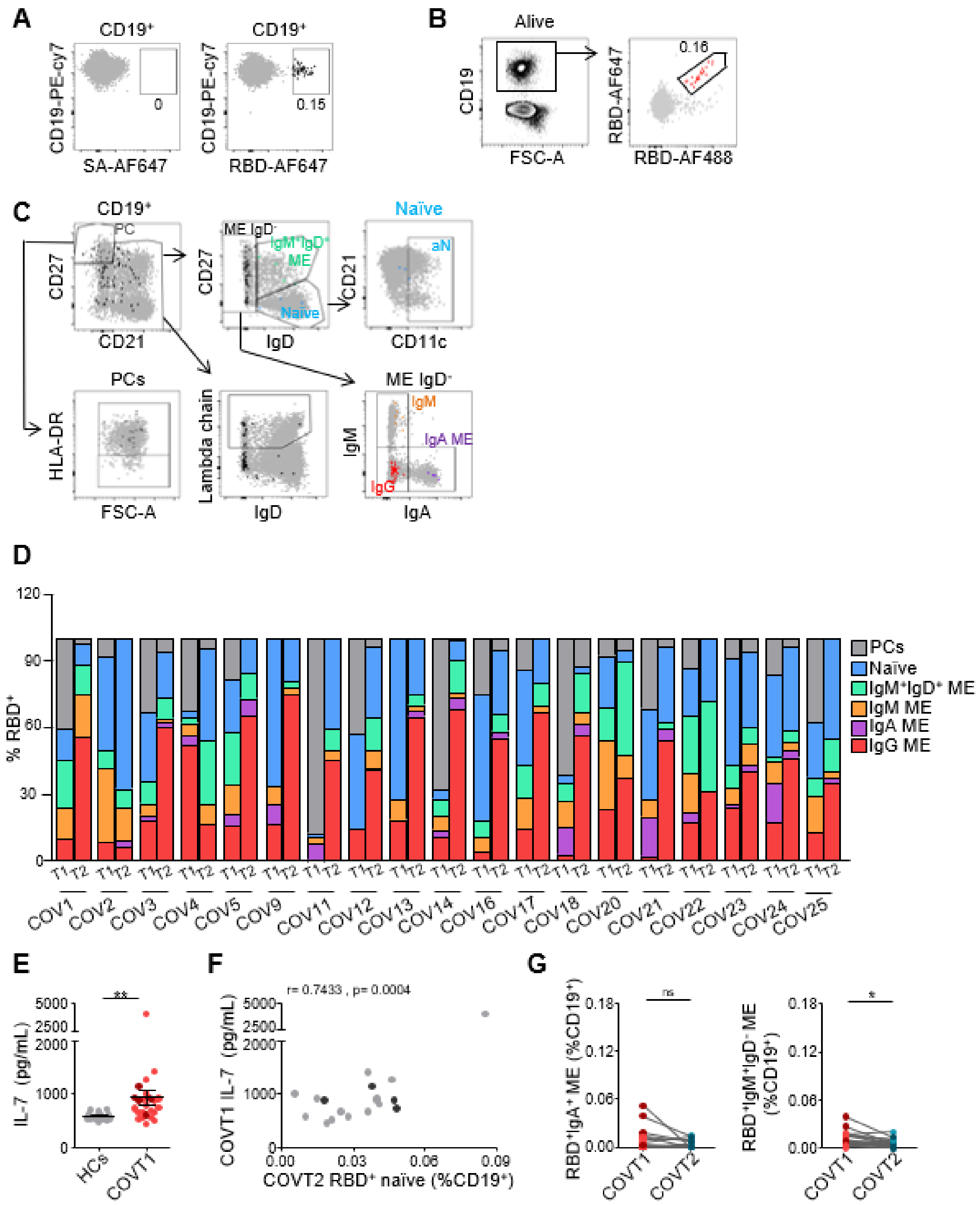


**Fig. S4 (related to Fig. 4). SARS-CoV-2 infection induces sustained RBD-specific IgG⁺ memory B cells.** (**A**) Flow cytometry plots showing staining without (left) and with (right) biotinylated RBD in total CD19⁺ cells represented by black large dots. Numbers indicate the percentage of RBD-specific cells within total CD19⁺ B cells. (**B**) Flow cytometry staining of CD19⁺ B cells from a representative convalescent COVID-19 patient using a double discrimination strategy through inclusion of two fluorescently labeled RBD probes in the same staining tube. Red large dots represent cells that are positive for both RBD-AF647 and RBD-AF488. Percentage indicates the proportion of

RBD-specific cells within total $CD19^+$ B cells. (**C**) Gating strategy used to define RBD-specific B cell populations: PCs ($CD19^+CD27^{++}CD21^-$), cells that produce lambda-expressing antibodies (non-PC $CD19^+lambda^+$), ME $IgD^-$ (non-PC $CD19^+IgD^-$), $IgM^+$ ME (non-PC $CD19^+IgD^-IgM^+$), $IgA^+$ ME (non-PC $CD19^+IgD^-IgA^+$), $IgG^+$ ME (non-PC $CD19^+IgD^-IgG^+$), $IgM^+IgD^+$ ME(non-PC $CD19^+CD27^+IgM^+IgD^+$), naïve (non-PC $CD19^+CD27^-IgD^+$) and $CD11c^+$ activated naïve (aN) B cells. RBD-specific cells are represented in colored large dots. (**D**) Relative percentage of B cell subsets within total $RBD^+CD19^+$ B cells in T1 and T2 for all patients analyzed. (**E**) Interleukin-7 (IL-7) levels in plasma of healthy controls (HCs; *n*=15)) and COVT1 (*n*=25) patients. (**F**) Pearson's correlation analysis of $RBD^+$ naïve B cells within $CD19^+$ cells in COVT2 and IL-7 plasma levels in COVT1. r stands for Pearson's correlation coefficient. (**G**) Paired analysis of the frequency of RBD-specific $IgA^+$ (left) and $IgM^+IgD^-$ (right) ME B cells within total $CD19^+$ in COVT1 and COVT2. (**E-G**) Data are presented as individual dots. Dark-colored dots show ICU patients. Two-tailed Mann-Whitney U test was performed to compare HCs with COVT1. Wilcoxon matched pairs test was performed to compare COVT1 with COVT2. Unless mentioned otherwise, HCs, *n*=15; COVT1, *n*=19; COVT2, *n*=19.

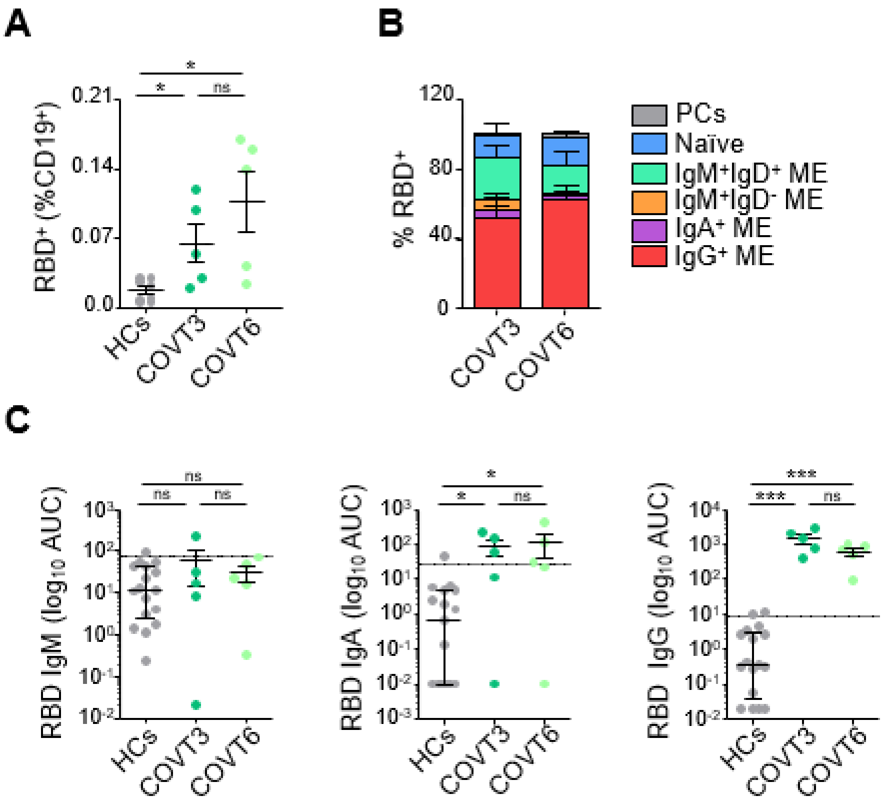


**Fig. S5 (related to Fig. 4). Long-term analysis of COVID-19 convalescent individuals suggest persistent expansion of RBD-specific IgG$^{+}$ memory B cells up to six months post-infection.** (**A**) Frequency of SARS-CoV-2 RBD-specific cells within total CD19$^{+}$ B cells, analyzed in healthy controls (HCs; *n*=8) and COVID-19 convalescent patients after 3 months (COVT3, *n*=5) and 6 months (COVT6, *n*=5) PSO. (**B**) Relative percentage of RBD-specific B cell populations among total RBD$^{+}$ B cells. Bars represent mean ± SEM. (**C**) Area under the curve (AUC) for each of the RBD-specific antibody classes analyzed from HCs, COVT3 and COVT6 sera samples. Sera from HCs were analyzed in parallel to establish negative threshold values defined as the HC AUC mean plus 2 times the standard deviation of the HC mean. Dashed line indicates negative threshold. Data are presented as individual dots. Bars represent mean ± SEM. Two-tailed Mann-Whitney U test was performed to compare HCs with COVT3 and HCs with COVT6. Wilcoxon matched pairs test was performed to compare COVT3 with COVT6 (*P < 0.05, **P < 0.01, and ***P < 0.001). Unless mentioned otherwise, HCs, *n*=17; COVT3, *n*=5; COVT6, *n*=5.

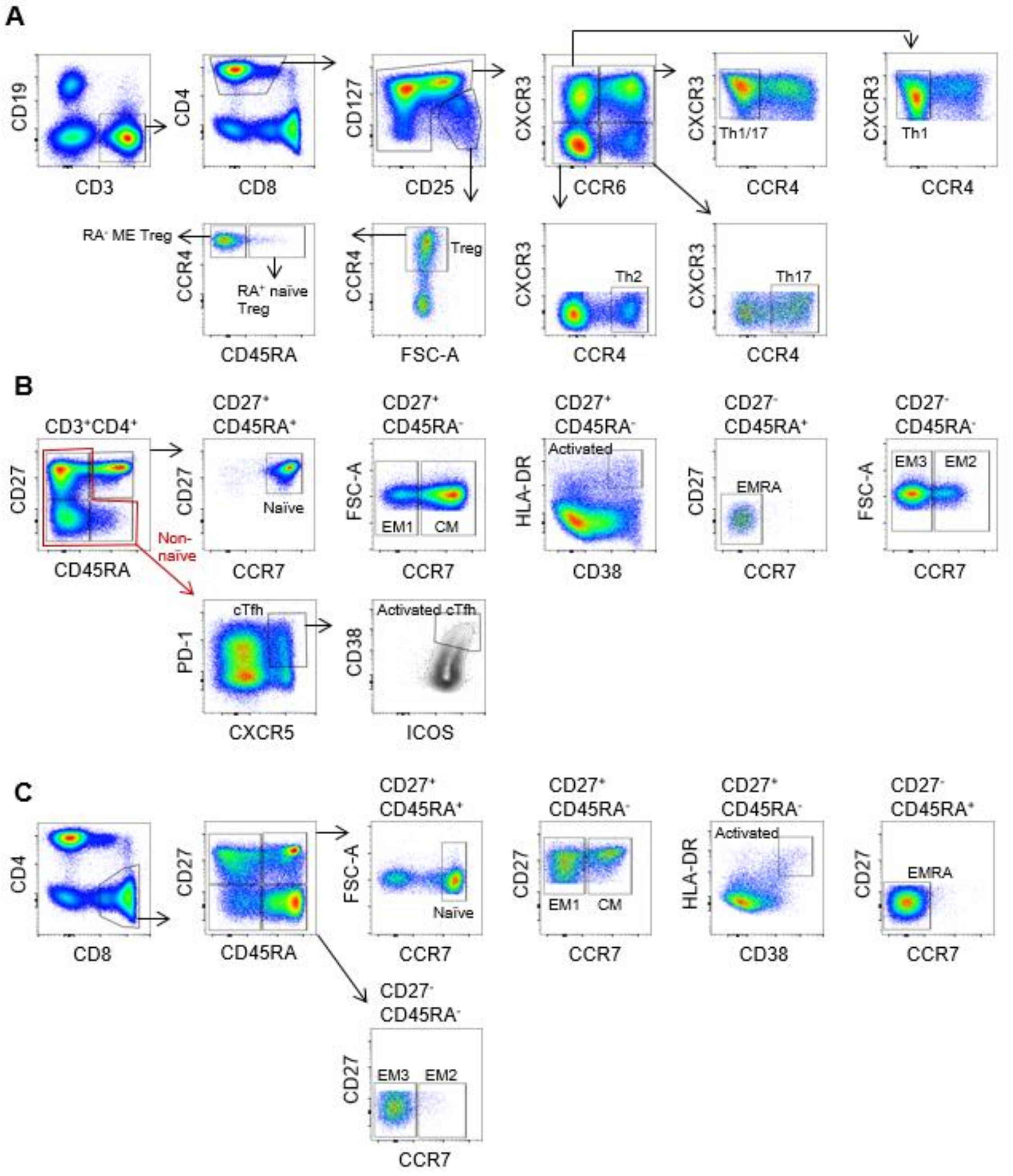


**Fig. S6 (related to Fig. 5). Gating strategy for major T cell populations.** (**A**) Gating strategy followed to define CD3$^+$CD4$^+$ T helper (Th) and regulatory T cells (Treg). T helper subsets were defined as non-Treg CXCR3$^+$CCR6$^+$CCR4$^-$ (Th1/17), non-Treg CXCR3$^+$CCR6$^-$CCR4$^-$ (Th1), non-Treg CXCR3$^-$CCR6$^-$CCR4$^+$ (Th2), and non-Treg CXCR3$^-$CCR6$^+$CCR4$^+$ (Th17). Treg subsets were defined as CD127$^-$CD25$^+$CCR4$^+$CD45RA$^-$ (RA$^-$ ME Treg) and CD127$^-$CD25$^+$CCR4$^+$CD45RA$^+$ (RA$^+$ naïve Treg). (**B**) Gating strategy followed to define CD3$^+$CD4$^+$ T cell subpopulations. Subsets were defined as CD27$^+$CD45RA$^+$CCR7$^+$ (naïve), CD27$^+$CD45RA$^-$CCR7$^+$ (CM, central memory), CD27$^+$CD45RA$^-$CCR7$^-$ (EM1, effector memory type 1), CD27$^+$CD45RA$^-$CD38$^+$HLA-DR$^+$ (activated), CD27$^-$CD45RA$^+$CCR7$^-$ (EMRA, effector memory RA$^+$), CD27$^-$CD45RA$^-$CCR7$^-$ (EM3, effector memory type 3), CD27$^-$CD45RA$^-$CCR7$^+$ (EM2, effector memory type 2), non-naïve PD-1$^+$CXCR5$^+$ (cTfh, circulating T follicular helper cells), and non-naïve PD-1$^+$CXCR5$^+$CD38$^+$ICOS$^+$ (activated cTfh). (**C**) Gating strategy followed to define CD3$^+$CD8$^+$ T cell subpopulations. Subsets were defined by CD27$^+$CD45RA$^+$CCR7$^+$ (naïve), CD27$^+$CD45RA$^-$CCR7$^+$ (CM, central memory), CD27$^+$CD45RA$^-$CCR7$^-$ (EM1, effector memory type 1), CD27$^+$CD45RA$^-$CD38$^+$HLA-DR$^+$ (activated), CD27$^-$CD45RA$^+$CCR7$^-$ (EMRA, effector memory RA$^+$), CD27$^-$CD45RA$^-$CCR7$^-$ (EM3, effector memory type 3), and CD27$^-$CD45RA$^-$CCR7$^+$ (EM2, effector memory type 2).

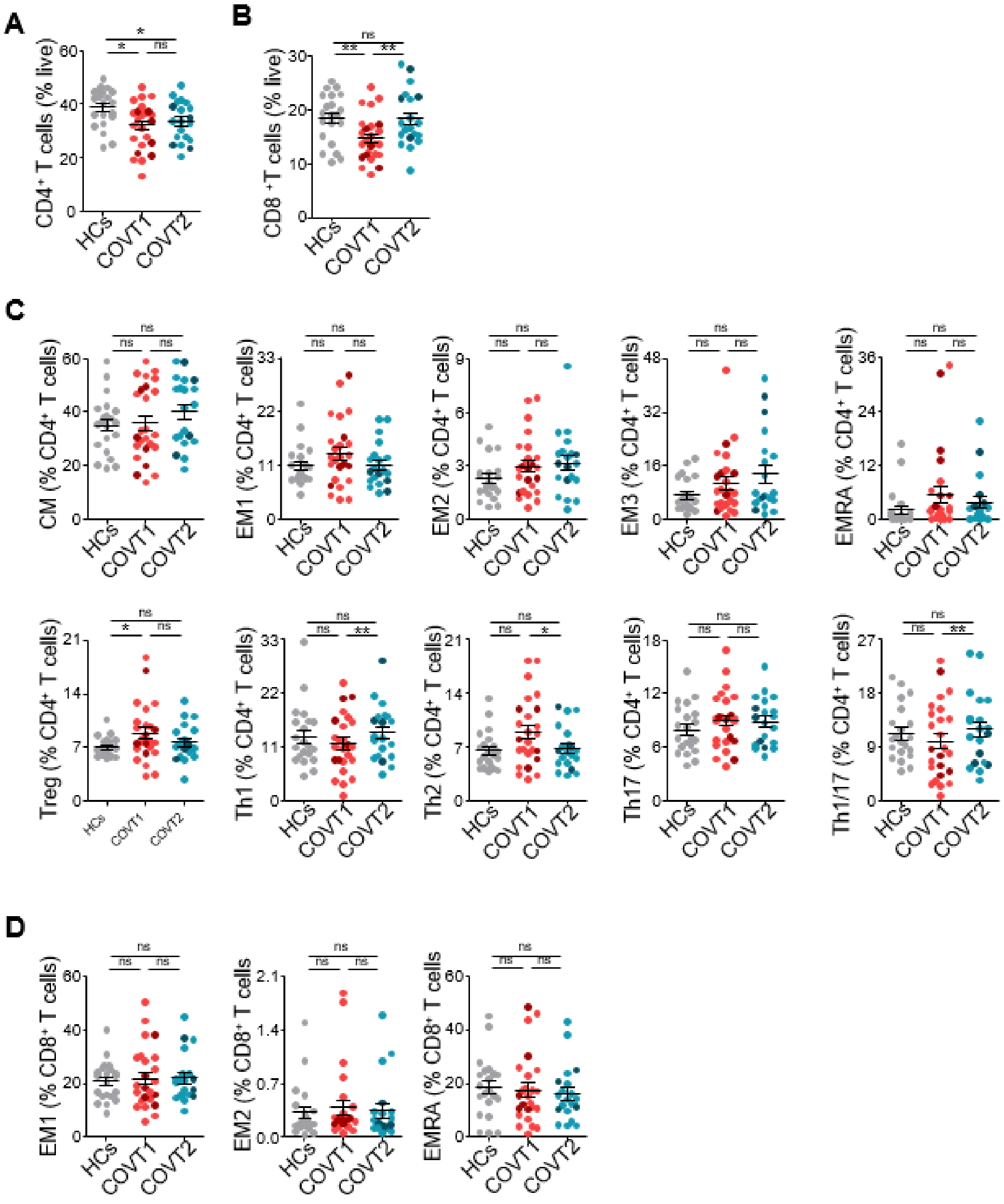


**Fig. S7 (related to Fig. 5). SARS-CoV-2 infection associates with changes in the relative frequency of $CD4^+$ T helper subsets.** (**A**) Frequency of $CD4^+$ T cells within total live cells. (**B**) Frequency of $CD8^+$ T cells within total live cells. (**C**) Frequencies of $CD4^+$ T cell subsets among total $CD4^+$ T cells, as depicted in **Fig. S6**. CM, central memory. EM, effector memory. EMRA, effector memory RA+. Treg, regulatory T cells. Th, T helper cells. (**D**) Frequencies of $CD8^+$ T cell subsets among total $CD8^+$ T cells, as depicted in **Fig. S6.** Data are presented as individual dots. Dark-colored dots show ICU patients. Bars represent mean ± SEM. Two-tailed Mann-Whitney U test was performed to compare healthy controls (HCs) with COVT1 and HCs with COVT2. Wilcoxon matched pairs test was performed to compare COVT1 with COVT2 (*P < 0.05, **P < 0.01, and ***P < 0.001). HCs, *n*=21; COVT1, *n*=25; COVT2, *n*=20.

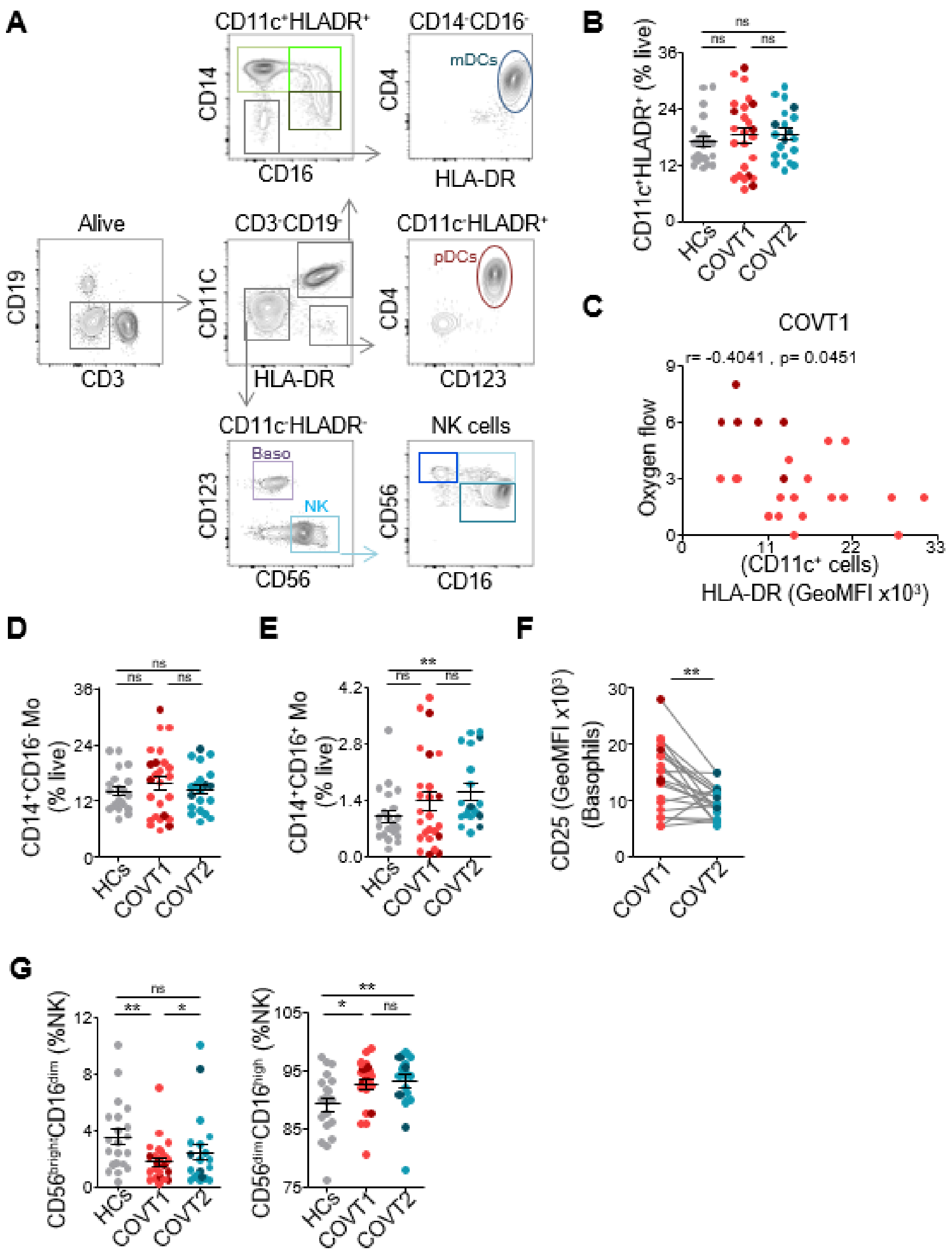


**Fig. S8 (related to Fig. 6). SARS-CoV-2 infection induces long-lasting changes in the innate immune compartment.** (**A**) Gating strategy used to define plasmacytoid dendritic cells (pDCs), myeloid dendritic cells (mDCs), basophils (baso), natural killer (NK) cells and monocyte (Mo) subsets. (**B**) Frequency of CD11c$^+$HLA-DR$^+$ myeloid cells within live PBMCs. (**C**) Spearman correlation analysis of oxygen flow in COVT1 plotted against GeoMFI of HLA-DR expression in CD19$^-$CD3$^-$CD11c$^+$ myeloid cells in COVT1. The oxygen flow ranges from 0, meaning no need for supplemental oxygen, to 8, meaning the need for orotracheal intubation and mechanical ventilation. r stands for Spearman's rank-order correlation. (**D**) Frequencies of CD14$^+$CD16$^-$ conventional and

(**E**) $CD14^+CD16^+$ intermediate Mo within live cells. (**F**) Paired analysis of CD25 GeoMFI in basophils from COVT1 (*n*=20) and COVT2 (*n*=20) samples. (**G**) Frequencies of NK cell subsets from total NK cells. Data are presented as individual dots. Dark-colored dots show ICU patients. Bars represent mean ± SEM. Two-tailed Mann-Whitney U test was performed to compare healthy controls (HCs) with COVT1 and HCs with COVT2. Wilcoxon matched pairs test was performed to compare COVT1 with COVT2 (*$P < 0.05$, **$P < 0.01$, and ***$P < 0.001$). Unless mentioned otherwise, HCs, *n*=21; COVT1, *n*=25; COVT2, *n*=20.

| VARIABLE | COVID-19 patients | HCs |
|---|---|---|
| Number of participants | 25 | 21 |
| **Demographics** | | |
| Age (years) | 51 [26-75] | 50 [31-64] |
| Sex (F/M) | 12 (48%) / 13 (52%) | 16 (76%) / 5 (24%) |
| **Disease characteristics** | | |
| Number of days from symptom onset to COVT1 | 11 [8-14] | NA |
| Days of hospitalization | 16 [6-90] | NA |
| NIH Ordinal Severity Score | 4 [2-6] | NA |
| Pulmonary Affection Severity Score | 5 [2-7] | NA |
| Oxigen Flow | 3 [0-8] | NA |
| ICU (yes/no) | 6 (24%) / 19 (76%) | NA |
| Fibrosis (yes/no) | 4 (16%) / 21 (84%) | NA |
| Intestinal symptoms (yes/no) | 9 (36%) / 16 (64%) | NA |
| Persistent Symptoms (yes/no) | 7 (28%) / 18 (72%) | NA |
| Number of days from symptom onset to COVT2 | 70 [59-84] | NA |
| **Laboratory clinical data** | | |
| D-dimer (µg/L FEU) | 1201 [350-6430] | NA |
| Ferritin (ng/mL) | 819 [49-3665] | NA |
| Interleukin 6 (pg/mL) | 168.2 [1.5-2459.0] | NA |
| Lactate dehydrogenase (UI/L) | 298 [142-591] | NA |
| C reactive protein (mg/dL) | 6.3 [0.4-18.8] | NA |
| Hemoglobin (g/dL) | 13.2 [10.5-15.9] | NA |
| Leucocytes (U/µL) | 7808 [4010-13600] | NA |
| Lymphocytes T (U/µL) | 1706 [720-3720] | NA |
| Platelets (U/µL) | 300160 [64000-523000] | NA |

**Table S1. Demographic and clinical data of patients included in the study**. Median and range are shown for continuous variables in demographics, disease characteristics and laboratory clinical data. Count and proportion are shown for categorical variable modalities. NIH ordinal severity score for hospitalized patients ranges from not requiring supplemental oxygen/no longer requiring ongoing medical care (6), not requiring supplemental oxygen/requiring ongoing medical care (5), requiring supplemental oxygen (4), on non-invasive ventilation or high flow oxygen devices (3), on invasive mechanical ventilation or extracorporeal membrane oxygenation (ECMO; 2), and death (1). Pulmonary affection severity score ranges from room air (RA; 7), nasal cannula (NC; 6), high flow nasal cannula/non-invasive ventilation (HFNC-NIV; 5), mild acute respiratory distress syndrome (ARDS; 4), moderate ARDS (3), severe ARDS (2), and severe ARDS with extracorporeal membrane oxygenation (ECMO; 1). Oxygen flow ranges from not requiring supplemental oxygen (0), requiring nasal cannula providing oxygen at 2 L/min and 28% concentration (1), requiring nasal cannula at 3 L/min and 32% (2), requiring nasal cannula 4 L/min and 35% (3), requiring Venturi mask that provides 50% oxygen concentration (4), requiring a non-rebreather mask that provides 70% oxygen concentration (5), requiring nasal high flow oxygen therapy (6), requiring non-invasive mechanical ventilation (7), and requiring orotracheal intubation and mechanical ventilation (8). NA = Not Applicable.

| **Antibodies MIX 1** | **Source** | **Reference** |
|---|---|---|
| Anti-human CD10 BV412 (clone: HI10a) | BioLegend | Cat# 312217; RRID:AB_10899409 |
| Anti-human CD19 Pacific Blue (clone: HIB19) | BioLegend | Cat# 302224; RRID:AB_493653 |
| Anti-human IgM Brilliant Violet 605 (clone: MHM-88) | BioLegend | Cat# 314524; RRID:AB_2562374 |
| Anti-human CD27 Brilliant Violet 650 (clone: O323) | BioLegend | Cat# 302827; RRID:AB_11124941 |
| Anti-human CD21 Brilliant Violet 785 (clone: B-ly4) | BD Biosciences | Cat# 740969; RRID:AB_2740594 |
| Anti-human IgG2 FITC (clone: SAG2) | Cytognos | Cat# CYT-IGG2F |
| Anti-human IgG3 FITC (clone: SAG3) | Cytognos | Cat# CYT-IGG3F |
| Anti-human IgA1 PerCP/Cy5.5 (clone: SAA1) | Cytognos | Cat# CYT-IGA1C |
| Anti-human IgA2 PerCP/Cy5.5 (clone: SAA2) | Cytognos | Cat# CYT-IGA2C2 |
| Anti-human IgG1 PE (clone: SAG1) | Cytognos | Cat# CYT-IGG1PE |
| Anti-human IgG2 PE (clone: SAG2) | Cytognos | Cat# CYT-IGG2PE |
| Anti-human IgD PE-CF594 (clone: IA6-2) | BD Biosciences | Cat# 562540; RRID:AB_11153129 |
| Anti-human IgG4 APC (clone: SAG4) | Cytognos | Cat# CYT-IGG4AP; RRID:AB_2876879 |
| Anti-human IgA1 APC (clone: SAA1) | Cytognos | Cat# CYT-IGA1AP |
| Anti-human HLA-DR Alexa Fluor 700 (clone: L243) | BioLegend | Cat# 307626; RRID:AB_493771 |
| Anti-human CD38 APC Fire 810 (clone: HB-7) | BioLegend | Cat# 356643; RRID:AB_2860936 |
| Anti-human CD24 PE/Cy7 (clone: ML5) | BioLegend | Cat# 311120; RRID:AB_2259843 |
| Anti-human CD43 Brilliant Violet 711 (clone: 1G10) | BD Biosciences | Cat# 743614; RRID:AB_2741624 |
| Anti-human CD11c APC/Cy7 (clone: Bu15) | BioLegend | Cat# 337217; RRID:AB_10661724 |
| Anti-human CXCR3 PE/Cy5 (clone: 1C6/CXCR3) | BD Biosciences | Cat# 561731; RRID:AB_10892799 |
| Anti-human CXCR4 Brilliant Violet 480 (clone: 12G5) | BD Biosciences | Cat# 746621; RRID:AB_2743901 |
| Anti-human CD45RB Alexa Fluor 594 (clone: MEM-55) | BioLegend | Cat# 310207; RRID:AB_2734281 |

**Table S2. Antibodies used for phenotypical characterization of B cell subsets by high-dimensional flow cytometry**. List of antibodies, source and reference used for the characterization of B cell subsets by high-dimensional flow cytometry. This antibody cocktail is referred to as MIX 1 in Materials and Methods section.

| Antibodies MIX 2 | Source | Reference |
|---|---|---|
| Anti-human CD19 PE/Cy7 (clone: HIB19) | BioLegend | Cat# 302216; RRID:AB_314246 |
| Anti-human IgM Brilliant Violet 605 (clone: MHM-88) | BioLegend | Cat# 314524; RRID:AB_2562374 |
| Anti-human IgD PE-CF594 (clone: IA6-2) | BD Biosciences | Cat# 562540; RRID:AB_11153129 |
| Anti-human HLA-DR Alexa Fluor 700 (clone: L243) | BioLegend | Cat# 307626; RRID:AB_493771 |
| Anti-human CD11c APC/Cy7 (clone: Bu15) | BioLegend | Cat# 337217; RRID:AB_10661724 |
| Anti-human CD27 PE (clone: O323) | eBioscience | Cat# 12-0279-42; RRID:AB_10718394 |
| Anti-human CD21 PE/Cy5 (clone: B-Iy4) | BD Biosciences | Cat# 551064; RRID:AB_394028 |
| Anti-human IgA Fc FITC | Invitrogen | Cat# H14101; RRID:AB_2536555 |
| Anti-human Ig light chain λ PerCP/Cy5.5 (clone: MHL-38) | BioLegend | Cat# 316617; RRID:AB_2561511 |
| **Alternative MIX 2 double staining strategy** | **Source** | **Reference** |
| Anti-human CD19 PE/Cy7 (clone: HIB19) | BioLegend | Cat# 302216; RRID:AB_314246 |
| Anti-human IgM Brilliant Violet 605 (clone: MHM-88) | BioLegend | Cat# 314524; RRID:AB_2562374 |
| Anti-human IgD PE-CF594 (clone: IA6-2) | BD Biosciences | Cat# 562540; RRID:AB_11153129 |
| Anti-human HLA-DR Alexa Fluor 700 (clone: L243) | BioLegend | Cat# 307626; RRID:AB_493771 |
| Anti-human CD11c APC/Cy7 (clone: Bu15) | BioLegend | Cat# 337217; RRID:AB_10661724 |
| Anti-human CD27 PE (clone: O323) | eBioscience | Cat# 12-0279-42; RRID:AB_10718394 |
| Anti-human CD21 PE/Cy5 (clone: B-Iy4) | BD Biosciences | Cat# 551064; RRID:AB_394028 |
| Anti-human IgA VioGreen (clone: IS11-8E10) | Miltenyi Biotec | Cat# 130-114-007; RRID:AB_2734098 |
| Anti-human Ig light chain λ PerCP/Cy5.5 (clone: MHL-38) | BioLegend | Cat# 316617; RRID:AB_2561511 |

**Table S3. Antibodies used for flow cytometric detection and phenotypical characterization of SARS-CoV-2-RBD-specific B cells**. List of antibodies, source and reference used for the characterization of RBD-specific B cell subsets by flow cytometry using a single discrimination strategy (upper panel) or double discrimination strategy (bottom panel). This antibody cocktail is referred to as MIX 2 in Materials and Methods section.

| Antibodies MIX 3 | Source | Reference |
|---|---|---|
| Anti-human CCR7 Brilliant Violet 421 (clone: G043H7) | BioLegend | Cat# 353208; RRID:AB_11203894 |
| Anti-human CCR6 PE/Dazzle 594 (clone: G034E3) | BioLegend | Cat# 353430; RRID:AB_2564233 |
| Anti-human CD16 Pacific Blue (clone: 3G8) | BioLegend | Cat# 302032; RRID:AB_2104003 |
| Anti-human CD3 Brilliant Violet 510 (clone: OKT3) | BioLegend | Cat# 317332; RRID:AB_2561943 |
| Anti-human CD8a Brilliant Violet 570 (clone: RPA-T8) | BioLegend | Cat# 301037; RRID:AB_10933259 |
| Anti-human CXCR5 Brilliant Violet 605 (clone: RF8B2) | BD Biosciences | Cat# 740379; RRID:AB_2740110 |
| Anti-human CD11c Brilliant Violet 650 (clone: B-ly6) | BD Biosciences | Cat# 563403; RRID:AB_2732048 |
| Anti-human CD123 Brilliant Violet 711 (clone: 6H6) | BioLegend | Cat# 306030; RRID:AB_2566354 |
| Anti-human CD56 Brilliant Violet 750 (clone: NCAM16.2) | BD Biosciences | Cat# 747068; RRID:AB_2871824 |
| Anti-human PD1 Brilliant Violet 785 (clone: EH12.2H7) | BioLegend | Cat# 329929; RRID:AB_11218984 |
| Anti-human CD14 Spark Blue 550 (clone: 63D3) | BioLegend | Cat# 367148; RRID:AB_2832724 |
| Anti-human CD45ra FITC (clone: HI100) | Thermo Fisher Scientific | Cat# 11-0458-73; RRID:AB_465072 |
| Anti-human HLADR APC/Fire 750 (clone: L243) | BioLegend | Cat# 307658; RRID:AB_2572101 |
| Anti-human ICOS PE (clone: C398.4A) | BioLegend | Cat# 313507; RRID:AB_416331 |
| Anti-human CD4 cFluor YG584 (clone: SK3) | Cytek | Cat# R7-20041-100T |
| Anti-human CXCR3 PE/Cy5 (clone: 1C6/CXCR3) | BD Biosciences | Cat# 561731; RRID:AB_10892799 |
| Anti-human CD25 PE/Alexa Fluor 700 (clone: CD25-3G10) | Thermo Fisher Scientific | Cat# MHCD2524; RRID:AB_2539740 |
| Anti-human CD19 PE/Cy7 (clone: HIB19) | BioLegend | Cat# 302216; RRID:AB_314246 |
| Anti-human CD27 APC (clone: M-T271) | BD Biosciences | Cat# 561786; RRID:AB_10896653 |
| Anti-human CCR4 Alexa Fluor 647 (clone: L291H4) | BioLegend | Cat# 359403; RRID:AB_2562388 |
| Anti-human CD127 APC/R700 (clone: HIL-7R-M21) | BD Biosciences | Cat# 565185; RRID:AB_2739099 |
| Anti-human CD38 APC Fire 810 (clone: HB-7) | BioLegend | Cat# 356643; RRID:AB_2860936 |

**Table S4. Antibodies used for phenotypical characterization of other circulating lymphoid and myeloid populations by high-dimensional flow cytometry**. List of antibodies, source and reference used for the characterization of T and innate immune cell subsets by high-dimensional flow cytometry. This antibody cocktail is referred to as MIX 3 in Materials and Methods section.

**Data File S1. Raw data of SARS-CoV-2-specific antibody titers and selected immune parameters used to perform pairwise correlations and PCA (provided as a separate Excel file)**. Raw data including virus-specific antibody titers measured by ELISA and selected immune parameters identified by high-dimensional flow cytometry used to perform pairwise correlations and PCA. NA = Not Applicable.
